# Evolution of synchronous bilateral breast cancers provide insights into interactions between host, tumor and immunity

**DOI:** 10.1101/2021.12.28.21267674

**Authors:** Anne-Sophie Hamy, Judith Abecassis, Lauren Darrigues, Cecile Laurent, François Zaccarini, Benjamin Sadacca, Myriam Delomenie, Enora Laas, Odette Mariani, Thanh Lam, Beatriz Grandal, Marick Lae, Ivan Bieche, Sophie Vacher, Jean-Yves Pierga, Etienne Brain, Celine Vallot, Judicael Hotton, Wilfrid Richer, Joshua Waterfall, Fabien Reyal

## Abstract

Synchronous bilateral breast cancer (sBBC) occurs after both breasts have been affected by the same germline genetics, reproductive life factors and environmental exposures for decades. It represents an opportunity to decipher the complex interplay between host, tumor, immune system and response to neoadjuvant chemotherapy (NAC). On a cohort of 17575 BCs treated between 2005 and 2012, sBBCs (n=404) were associated with less aggressive proliferative patterns and higher rates of luminal breast cancers (BCs) when compared with unilateral BCs (n=17171). The left and right tumors were concordant for the majority of clinical and pathological features. Tumor pairs of concordant BC subtype were more frequent than pairs of discordant BC subtype, with notably a particularly high frequency of pairs of luminal BCs. Intriguingly, both the levels of tumor infiltrating lymphocytes (TILs) and the response to NAC were modified by the subtype of the contralateral tumors. Whole exome sequencing and RNAseq analyses revealed that left and right tumors were independent from a somatic mutation and transcriptomic point of view, while primary tumors (PT) before NAC and specimens with residual disease (RD) after NAC were more closely related. The analysis of the TCR repertoire identified very little overlap between patients, while common clones were shared in bilateral tumors within each patient. After NAC, the TCR repertoire of RD was enriched and expanded with clones edited by the contralateral PT.

## Introduction

Bilateral breast cancers (BBCs) represent 2% to 11% of breast cancers ^1–3^ and their incidence is increasing due to advances in breast cancer imaging ^4^. This entity includes both synchronous bilateral breast cancers (sBBCs) and metachronous bilateral breast cancer (mBBCs), according to the length of time between the two diagnoses, with typical cut-offs ranging from 3 to 12 months after diagnosis of the first tumor. sBBCs are associated with a poorer survival than unilateral cancer in several studies ^2,5,6^. In a study of 123,757 women with a primary BC diagnosed from 1970 to 2000, the cumulative breast cancer specific mortality was 45% for women with sBBC and 33% (p<0.001) in case of unilateral breast cancer ^2^. Beyond familial history of breast cancer, which is more frequent in women with BBC, neither synchronous nor metachronous breast cancer is associated with strong genetic determinants and only 5% of patients with BBC carry *BRCA1* or *BRCA2* mutations ^5^. Hence, per the 2020 NCCN guidelines, bilaterality is not considered as a factor clinically indicating genetic testing, but it may be considered in patients diagnosed with BBC between the age of 50 and 65 years ^7^.

From the genomic point of view, several studies have investigated clonal relationships between BBCs, with the majority reaching the conclusion that most of – if not all - BBCs were independent events ^8–14^. These studies used several methods to address the question of the origin of BBCs, including X chromosome inactivation analysis, comparisons of allelic imbalance patterns ^10,12,15^, or the distribution of p53 mutations ^11,16^, karyotyping ^17^, and comparative genomic hybridization (CGH) ^14,18^. Many of these studies were based on technologies with limited resolution for assessing clonality. Recently, analyzing a targeted sequencing panel of 254 genes including all known driver genes in BC, Begg and colleagues^19^ investigated the clonality of 30 synchronous and 19 metachronous BBC pairs, and found that only 3 out of 49 pairs harbored strong molecular evidence supporting a clonal relationship between BBCs. However, only one pair of sBBCs was interpreted as clonally related (2 shared mutations out of 3 mutations identified), leaving the question of the independence between sBBCs still open.

The immune microenvironment and especially the role of tumor-infiltrating lymphocytes (TILs) in BC has been studied extensively in the last decade. The drivers of BC immunosurveillance derive from both (i) tumor-intrinsic characteristics; and/or (ii) extrinsic factors related to the host or the environment. Among endogenous tumor characteristics, molecular features (BC subtype, proliferative patterns), expression of human leukocyte antigen (HLA)-class I, tumor mutational burden ^20^, and activation of particular cellular signaling pathways ^21^ have been found to be associated with immune infiltration. Extrinsic factors including host characteristics (gender ^22^, age ^23^, body mass index), environment (tobacco, alcohol), nutritional factors, diet, commensal microbiota, physical activity, and hormonal exposure have been studied less extensively. Even if our understanding of the immune microenvironment has improved, it remains unclear to what extent anti-tumor immunity is driven by the tumor, by the host, or the interaction between the host and the tumor.

Neoadjuvant chemotherapy (NAC) is currently administered to patients with locally advanced breast cancers. Beyond increasing the rate of breast-conserving surgery, it serves as a test of *in vivo* chemo-sensitivity, and the analysis of residual tumor burden may help understanding resistance to treatment. Molecular BC subtypes and the density of tumor-infiltrating immune cells are both considered as important predictive and prognostic factors for optimal risk stratification and treatment individualization. Many studies have reported associations between high levels of TILs at diagnosis and better response to NAC ^24,25^, and better prognosis in both neoadjuvant and adjuvant chemotherapy, particularly for triple negative BCs (TNBC) and *HER2*-positive BC ^26^.

sBBCs occur after both breasts have been affected by the same germline genetics, reproductive life factors and environmental exposures for several decades. Two tumors arising concomitantly in a host mimic a model where (i) *extrinsic factors* are almost fully shared by the same host; (ii) *intrinsic factors* are specific to the tumor of each side; (iii) the *immune tumoral microenvironment* resulting of the interaction between the same host and two different tumors can be compared. In the case where these tumors are treated by NAC, sBBCs represent a unique opportunity to decipher the interactions between tumor, microenvironment, and response to treatment.

In this study, we describe on a large institutional cohort of sBBCs the relationships between factors related to host, tumor phenotypes, levels of TILs and response to NAC. In addition, we identified a rare resource of highly selected tumors, *i*.*e*., sBBCs treated by NAC, and performed whole exome sequencing (WES), copy number variation, and RNA sequencing (RNAseq) to draw a comprehensive analysis of somatic alterations, the immune microenvironment and the tumor evolution under treatment.

## Results

### Patient and tumor characteristics

Out of 17575 BC patients in our institutional clinical database, 404 patients had synchronous bilateral BC (2.3%) (FigS1). Patients with sBBC were slightly older at diagnosis (60.4 *versus* 58.2 years, *p*=0.001) and had a higher mean BMI (25.6 *versus* 24.7, *p*<0.001) than patients with unilateral breast cancer. A lower proportion of grade 3 (20.9% *versus* 29.4%, *p*<0.001), tumors of TNBC (7.7% *versus* 11.1%) or *HER2*-positive (6.1% *versus* 11.4%, *p*<0.001) subtype, or with lymphovascular invasion (21.2% *versus* 28.4%, *p*<0.001) were observed in bilateral tumors than in unilateral BCs (TableS1 and FigS2). ER and PR staining percentages were lower in unilateral than in bilateral BCs. Out of 313 patients with invasive sBBCs, most of the tumors were luminal (n=538, 87.6%), whereas TNBC (n=44, 7.2%) and *HER2*-positive BCs (n=32, 5.2%) were rare (TableS2). Overall, paired sBBC tumors shared more common characteristics than expected by chance (TableS3): the majority (84.7%) of the tumor pairs were concordant regarding clinical and pathological patterns, notably regarding BC subtype (Fig1A). In luminal tumors, ER and PR staining intensity of stained cells were lower when the pair of tumors was discordant than when the pair of tumors was concordant (Fig1B-C).

**Fig1:**
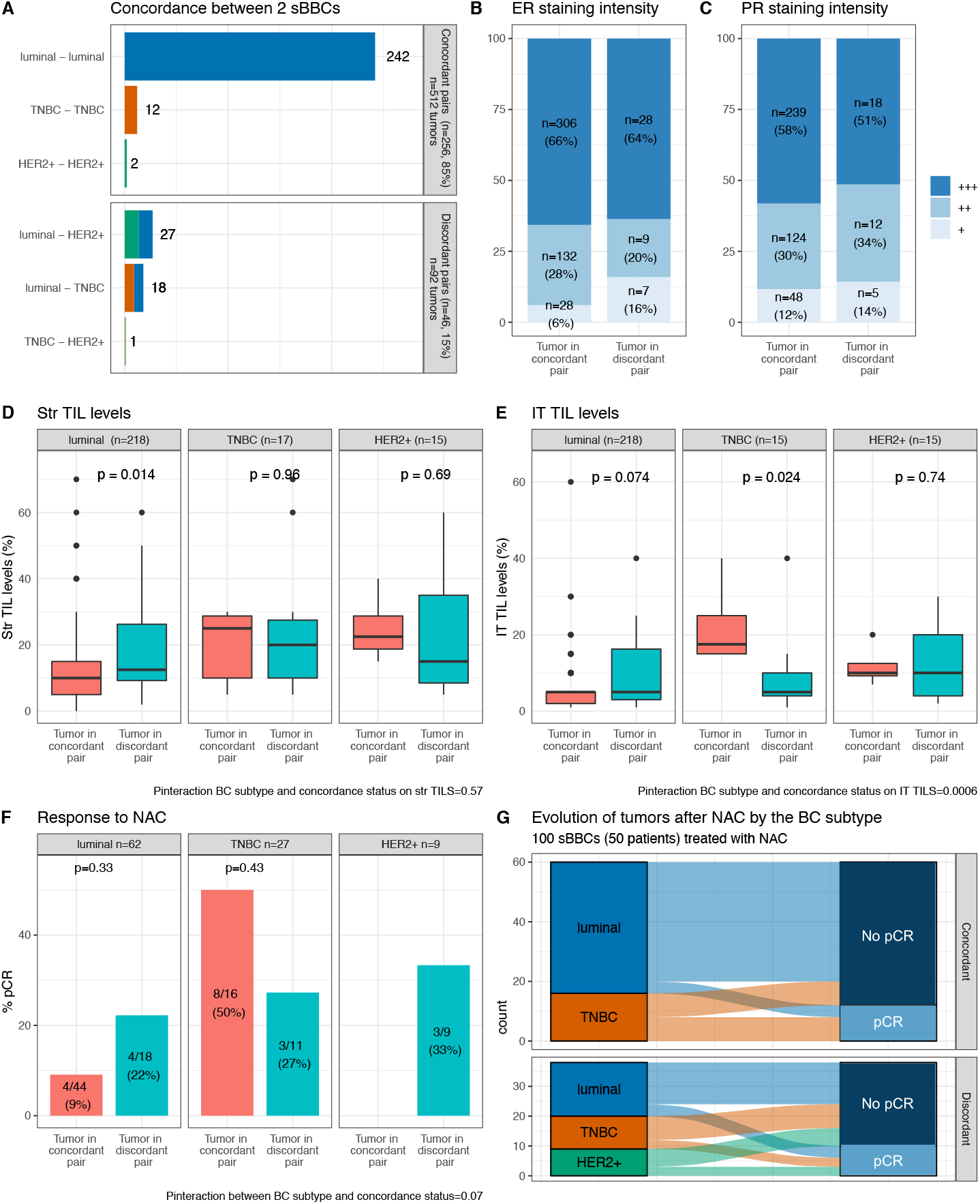
Tumor’s characteristics of the pairs of sBBCs included in the cohort. Tumor’s characteristics are based on the 302 pairs with concordance subtype available out of 313 pairs. The concordance refers to the status of both tumors within a pair of sBBCs regarding BC subtype, either of the same BC subtype (tumor in concordant pair), either of different BC subtypes (tumor in discordant pair). A. Repartition of the association of the BC subtypes within a pair of sBBCs according to the concordance or the discordance status of the pair; B. ER staining intensity of the index tumor by the concordance status of the pair it belongs to (luminal tumors only, n=510); C. PR staining intensity of the index tumor by the concordance status of the pair it belongs to (luminal tumors only, n=446); D. Str TIL levels of the index tumor by BC subtype and by the concordance status of the pair it belongs to; E. IT TIL levels of the index tumor by BC subtype and by the concordance status of the pair it belongs to; F. pCR rates of the index tumor by BC subtype and by the concordance status of the pair it belongs to; G. Evolution of the 100 tumors after NAC according to the BC subtype of the index tumor and by the concordance status of the pair of tumor. Abbreviations: BC, breast cancer; ER, estrogen receptor; IT TIL, intra tumoral tumor infiltrating lymphocytes. NAC, neoadjuvant chemotherapy; pCR, pathologic complete response; PR, progesterone receptor; sBBC, synchronous bilateral breast cancers; Str TIL, stromal tumor infiltrating lymphocytes; TNBC, triple negative breast cancer

### Baseline immune infiltration

Immune infiltration levels before treatment were available for 277 pairs of tumors. TIL levels were positively associated notably with a younger age, palpable, higher grade tumors, of no specific histological type, with a higher number of nodes involved and the presence of lymphovascular invasion (TableS4). TILs were higher in TNBC and *HER2*-positive BCs than in luminal tumors. Interestingly, the relationship between TIL levels and BC subtype showed a systemic effect, *i*.*e*., it was affected by the subtype of the contralateral tumor. In luminal BCs, stromal (Str) TIL levels were lower when the subtype of the contralateral tumor was concordant than when it was discordant, and the same trend was observed for intratumoral (IT) TILs (Fig1D-E). Conversely, in TNBCs, the IT TIL levels were lower when the subtype of the contralateral tumor was concordant than when it was discordant. The interaction test was highly significant (*P*_*interaction*_=0.0006), indicating that the impact of tumor subtype on IT immune infiltration was significantly modified by the concordance of the BC subtype of the tumor pair it belonged to. This suggests that TIL levels are not purely determined by local tumor microenvironment properties.

### Response to NAC

Out of 313 patients, 50 patients received neoadjuvant chemotherapy (NAC). Twenty two out of 100 tumors reached pathological complete response (pCR). In pairs with concordant tumor types, the pCR rate was higher for TNBC tumors than for luminal BC (50% *versus* 9%) (Fig1F-G). As was seen for TIL levels, these pCR rates showed a systemic effect when the contralateral tumor subtype was discordant. The pCR rate was higher in luminal BCs when the contralateral pair was of different subtype (9% *versus* 22%); while the opposite pattern was found in TNBCs (50% *versus* 27%), and the interaction test was significant (*P*_*interaction*_ =0.07), suggesting that the response to treatment was modified by the BC subtype of the contralateral tumor differentially in luminal and in TNBCs tumors.

### Genome-scale analyses

For 6 patients with sBBCs treated with NAC, frozen material of sufficient quality was available to perform tumor/normal whole exome sequencing (WES) and tumor RNA sequencing (RNAseq) in both left and right pre and post NAC samples (including one patient with a multicentric bilateral breast cancer) (FigS3, TableS5). Germline pathogenic mutations in BC predisposition genes were identified in four patients (*BRCA1*, n=2, *BRCA2*, n=2). Among the 14 primary tumors (PT), 9 were of luminal subtype and 5 were TNBCs. All patients received standard sequential anthracyclines-cyclophosphamide followed by taxanes. After NAC completion, 6 out of 14 tumors had residual disease (RD), while 8 tumors reached pCR.

#### Somatic single nucleotide and indel mutations

Twenty tumor samples (pre-NAC: n =14; post-NAC: n=6) were profiled by WES and RNAseq and distant juxta-tumor samples were used for germline WES sequencing in each patient. A median of 151.5 somatic mutations were detected per tumor (PT: 161.5 *versus* RD: 54), and a median of 3 mutations were annotated as potential drivers (splice site, nonsense, frameshift or non-synonymous SNVs in COSMIC census gene) in each sample (PT: 3.5 *versus* RD: 3) (TableS6). Tumors that reached a pCR had a similar median number of mutations (median 195 (driver: 5)) to those who had RD (median: 84 (driver: 2.5) and no difference was seen according to BC subtype (luminal: 160 (driver: 3) *versus* TNBC: 182 (driver: 4)).

The three most frequently mutated COSMIC genes were *TP53* (3 patients, 8 samples), *PTEN* (3 patients, 3 samples), and *ARID1A* (2 patients, 2 samples) (Fig2A). The majority of the remaining genes were mutated only in single patients, consistent with a high genetic diversity of mutations BC. No mutation was shared between the left and right side of the PTs from any patient, consistent with the contralateral tumors developing from independent clones (Fig2B). The majority of the mutations of a tumor were shared between a PT and its RD (median percentage of shared mutations in pairs of PT/RDs: 51.3%).

**Fig2:**
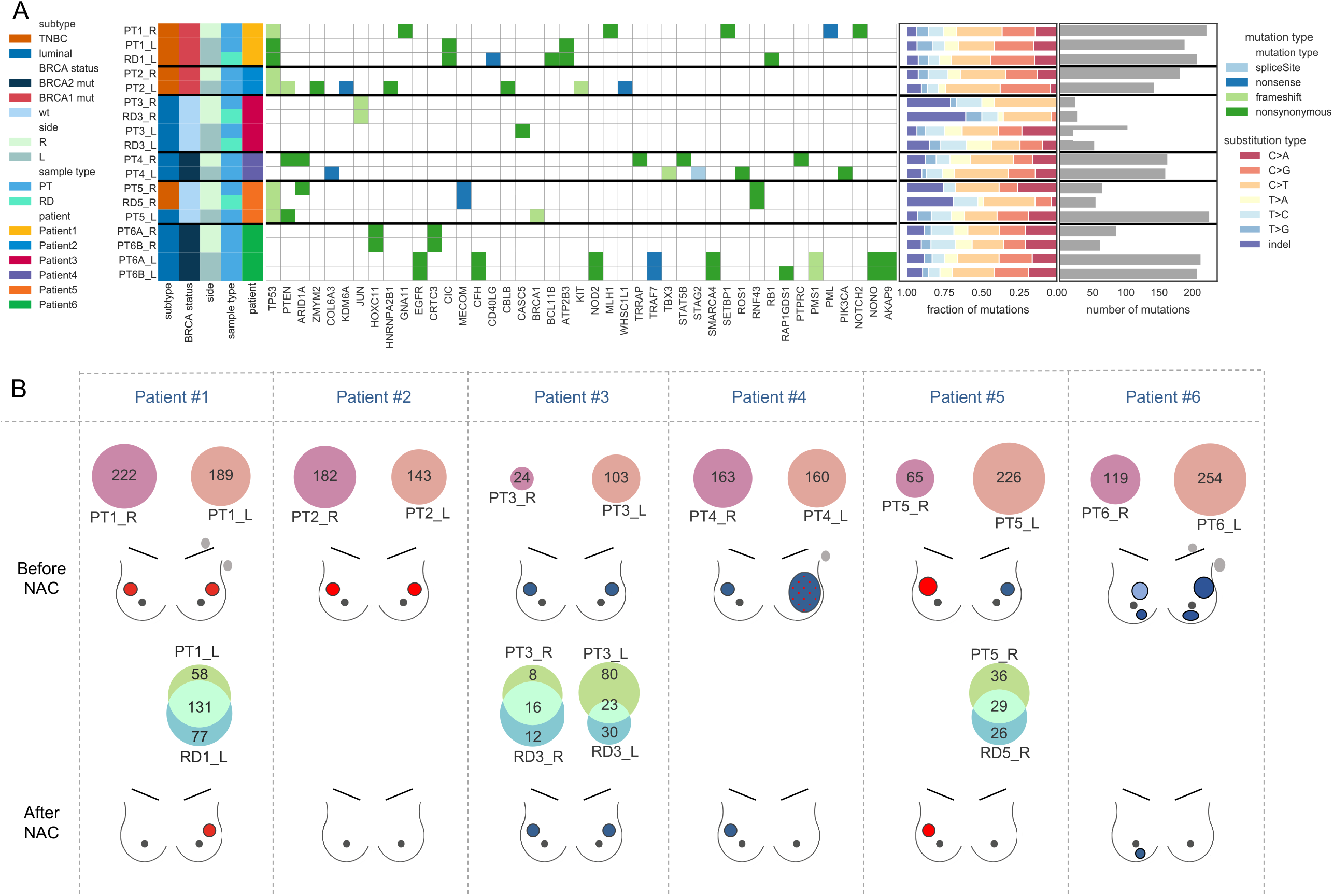
Tumor mutation profiles. The analyses are performed in the in left, right, pre and post-NAC samples of a cohort of 6 patients (20 samples) with next generation sequencing data available. A. Heatmap of somatic driver mutations (including missense, nonsense, and splicing) detected in 6 study patients and 20 samples. B. Venn diagrams showing the number of mutations shared between the left (pink) and the right (purple) primary tumors of a same patient; and shared (intersect turquoise) between the PT (light green) and the corresponding specimen after NAC (RD, residual disease, blue). The post NAC samples RD4_R and RD6B_R were discarded due to poor sample purity. Mutations from the 2 multicentric tumors of each side of patient #6 have been merged. Abbreviations: L, left; NAC, neoadjuvant chemotherapy; pCR, pathologic complete response; PT, primary tumor; R, right; RD, residual disease; TNBC, triple negative breast cancer

#### Copy number alterations (CNAs)

Copy number analysis of the WES profiles identified recurrent gains (8 out of 12 samples) at 1q, 8q, 17q and recurrent losses (over 50% of the samples) at 4p, 8p, 6q, 13q, 16q and in a lesser extent 1p (FigS4). Most of the alterations were not shared between the left and the right side (Fig3A), while most of the CNAs were common between PT and RD (Fig3B).

**Fig3:**
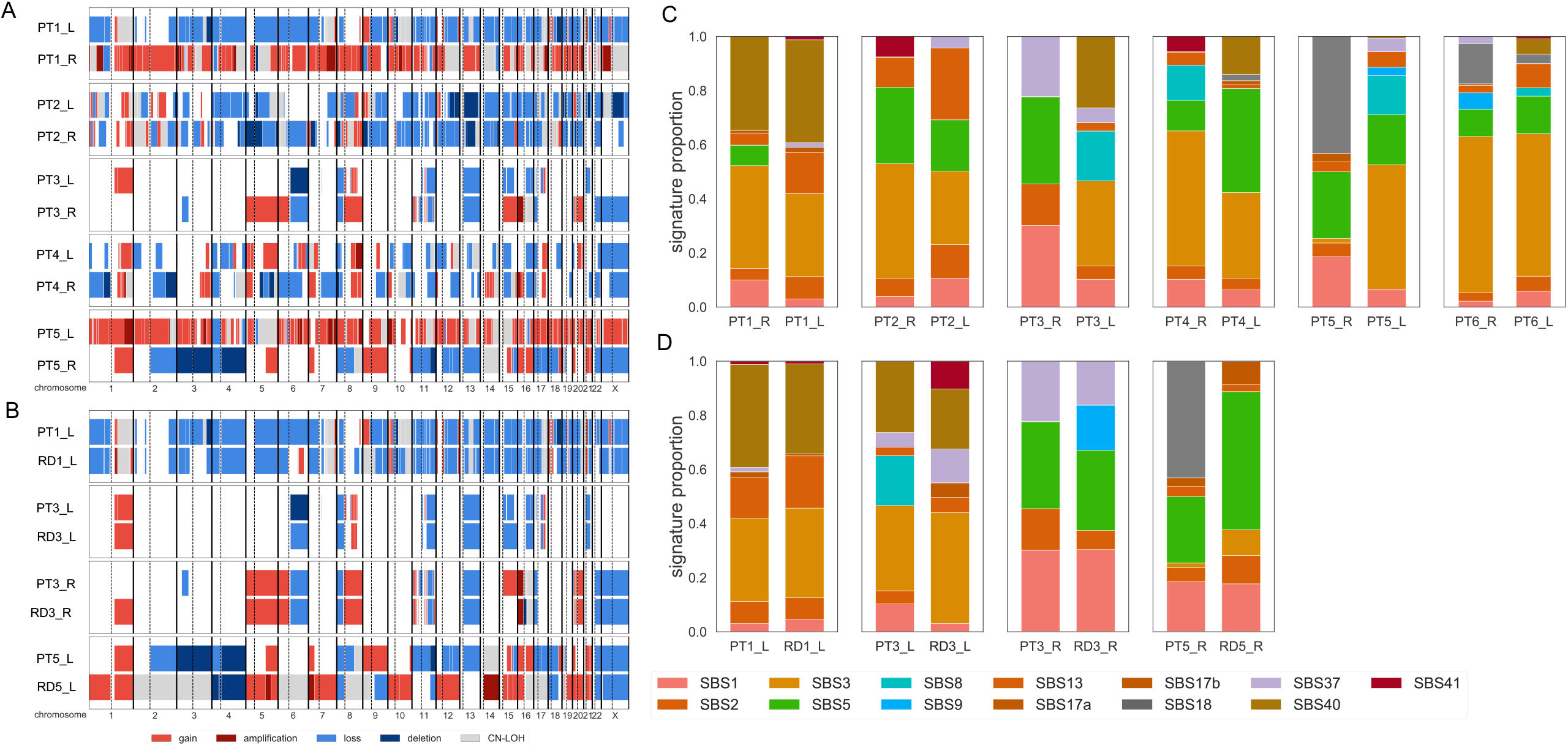
Copy number alterations (CNA) and mutational signatures profiles. The analyses are performed in the in left, right, pre and post-NAC samples of a cohort of 6 patients (20 samples) with next generation sequencing data available. Mutational signatures published by Alexandrov ^27^ are calculated using the deconstructSigs package. A. CNA are compared among the two sides of the primary tumor of the same patient; B. Among the primary tumor and its corresponding sample with RD after NAC. C. Mutational signatures are compared among the two sides of the primary tumor of the same patient (Mutations from the 2 multicentric tumors of each side of patient #6 have been merged.) and; D. Among the primary tumor and its corresponding sample with RD after NAC. Abbreviations: BC, breast cancer; CAN, copy number alterations; CH-LOH, copy-neutral loss of heterozygosity; chr, chromosome; L, left; NAC, neoadjuvant chemotherapy; R, right; RD, residual disease

#### Mutational signatures

We analyzed mutational signatures by deconvoluting the frequency of the 96 different possible trinucleotide substitutions against known signatures of mutation patterns ^27^. We found that the four most abundant base substitution signatures were signature 3 (34%), signature 5 (17%), and signature 40 (10%) (Fig3C). Signature 3 is known to be associated with *BRCA1/2* mutations (Alexandrov et al., 2020) and its contribution was high (54%) in all tumors from patients carrying *BRCA* germline mutations (Patient #1, #2, #4, #6) (Fig3C). Furthermore, signature 3 was also found in the left tumor of patient #5 who carried a somatic *BRCA1* mutation, while it was absent from the right-side tumor which did not show any *BRCA1-2* mutations. Signatures 5, 40 and 1 are commonly found in diverse tumor types and are associated with aging and were generally found at consistent levels between bilateral tumor pairs. Similarities regarding mutational processes were lower within the left and right side of the PTs than within pairs of PT-RDs (Fig3D).

#### Clonality and phylogenetic evolution

We determined the phylogenetic evolution between the germline profile of the patient to the left and right primary tumors and ultimately to the residual disease if present (Fig4A-E and supplemental results). The number of clones identified in each tumor sample ranged from 2 to 6. Genomic profiling found no common clones between bilateral primary tumors of the same patient, showing that these tumors arose through unrelated tumor evolution processes. Further illustrating this, left and right tumors showed distinct evolution regarding the homologous recombination deficiency (HRD), with the emergence of a somatic mutation on the *BRCA1* gene in only one out of the two tumors of a patient (Fig1E).

**Fig4:**
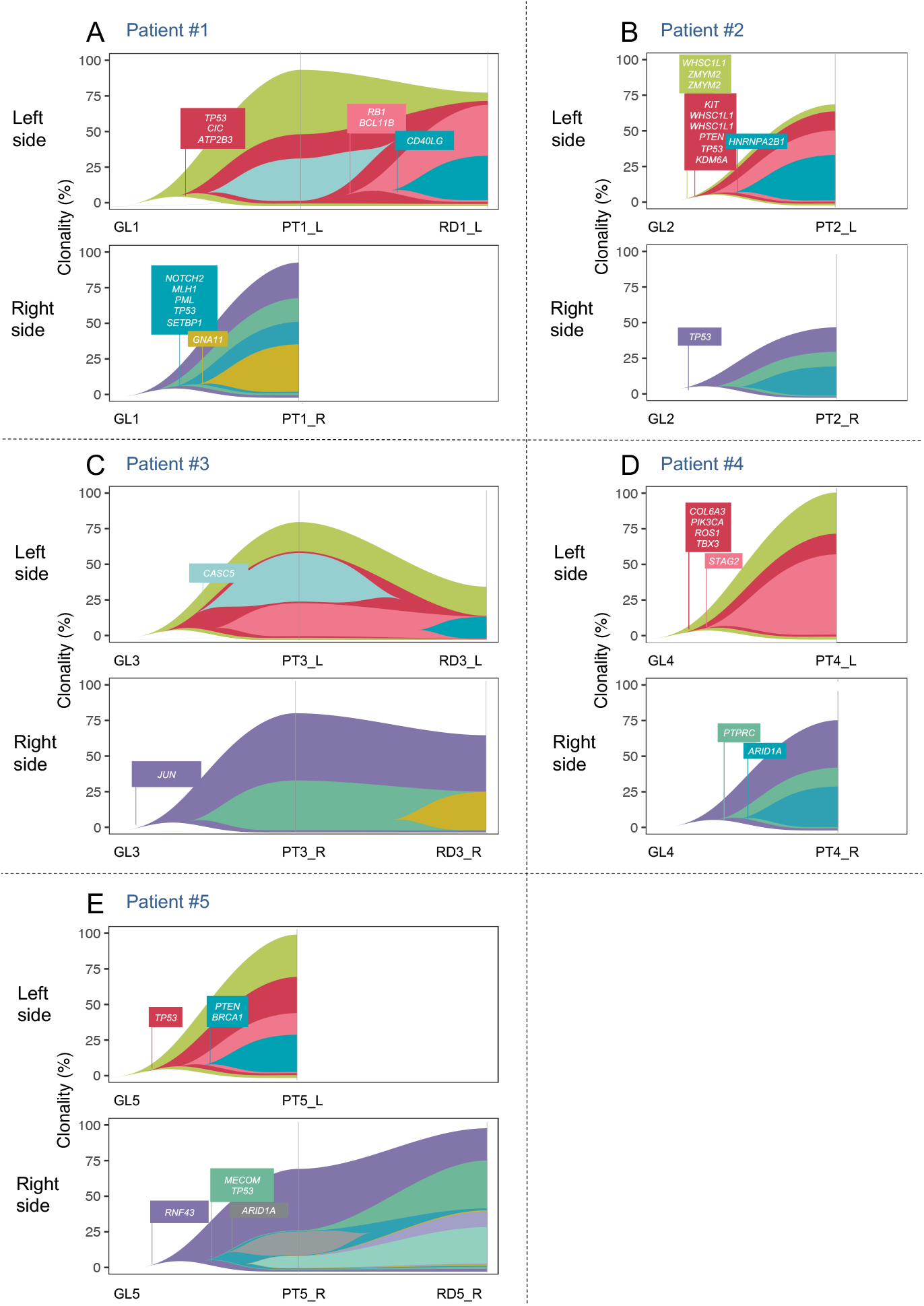
Fishplot retracing phylogeny between left, right primary tumors and corresponding residual disease. Each subfigure represents the evolution of the tumors of a given patient under NAC. The upper fish plot represents the evolution of the left tumor(s), the lower fishplot represents the evolution of the right tumor(s). Each fishplot displays the prevalence of subclones throughout treatment. Subclonal architecture was reconstructed with SuperFreq. Subclonal profiles show annotated common driver cancer genes.A. Both PTs were mutated for P53, but the genomic alteration was different on the left and the right side (right side: indel frameshift deletion position 7578213; left side: substitution C>T p.R175C missense substitution identified as pathogenic in Clinvar, and present in the RD); B. Both tumors were mutated for TP53 with 2 different mutations Both tumors were mutated for TP53 with 2 different mutations (right side: frameshift loss of a nucleotide position 7577558; E. TP53 was mutated on both sides (left side: frameshift deletion position 7578213, right side: frameshift deletion position 7579522); Abbreviations: NAC, neoadjuvant chemotherapy; PT, primary tumor; RD, residual disease.

For patient #3, for which we monitored evolution after NAC in both tumors in parallel, we did not observe any common clonal evolution after NAC. In all tumors, several clones disappeared under NAC (Fig4A and Fig4C, light turquoise clones; Fig4E grey clone), consistent with subclone specific responses to therapy. Finally, the majority of clones that emerged after NAC did not show identifiable additional genetic drivers, suggesting potential non genetic additional mechanisms could play a role in the tumor evolution under treatment. Altogether, these results suggest that left and right PTs are not clonally related and that their evolution under NAC does not converge to a common profile. Hence, RD is closer to its associated primary tumor than to its concomitant contralateral tumor.

#### Particular case of multicentric tumors

One patient of the six had a bilateral multicentric tumor (Patient #6) in the context of a *BRCA2* pathogenic germline mutation. While the left and the right tumors shared no common mutations, the two tumors from each side shared a majority of common genetic alterations at both the substitution (Fig5A) and copy number levels (Fig5B). Mutation signature analyses demonstrated a dominant genomic HRD footprint (signature 3, Fig5C). On each side, phylogenetic reconstruction clearly indicated that multicentric tumors were clonally related, with one tumors evoluting to a neighboor tumor through the extinction / emergence of particular subclones.

**Fig5:**
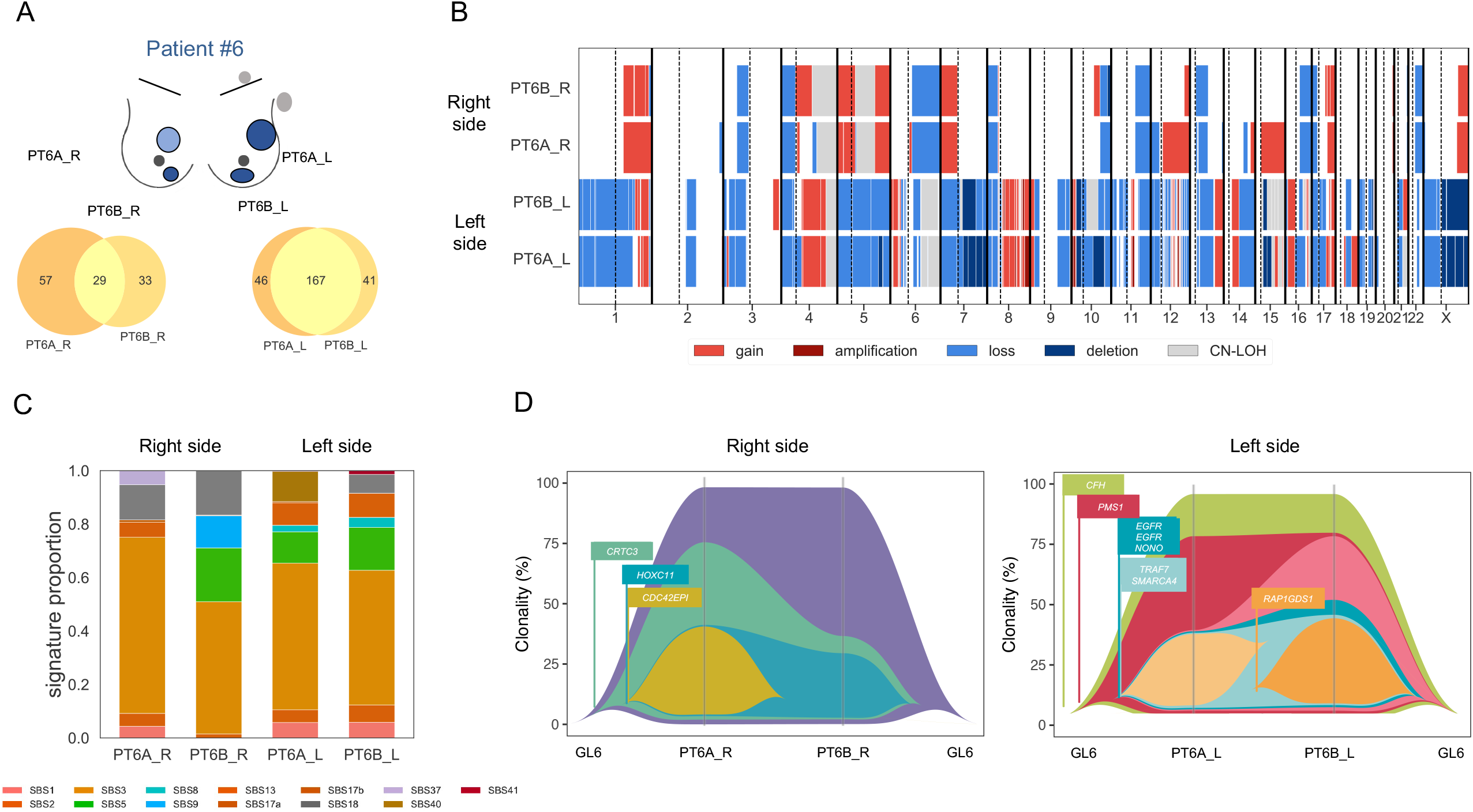
Genomic alterations in the patient with multicentric sBBCs. The analyses are performed in the in two left tumors (PT6A_L – PT6B_L), and in the 2 right tumors (PT6A_R – PT6B_R). The sample with residual disease (RD6B_R) was discarded due to poor sample purity. A. Tumor mutation profiles: Venn diagrams showing the number of mutations shared between the 2 primary tumors of each side (yellow intersect); B. Copy number alterations (CNA) profiles, compared among the two multicentric tumors of each side; C. Mutational signatures as from Alexandrov ^27^ compared among the two multicentric tumors of each side; D. Fishplot retracing the phylogeny between the two multicentric tumors of each side. Each fish plot can be interpreted from the left to the right, or from the right to the left, corresponding to the emergence or the extinction of a clone respectively. Abbreviations: L, left; PT, primary tumor; R, right; RD, residual disease; BC, breast cancer; CAN, copy number alterations; CH-LOH, copy-neutral loss of heterozygosity; chr, chromosome.

#### Transcriptomic alterations

##### Tumor clustering

We first performed unsupervised hierarchical clustering based on transcriptomic profile of the most variable genes. With the exception of tumor PT_5L, the sample clustering first split by BC subtype (luminal and TNBCs) (FigS5), while gene clustering split the 2846 genes into four main clusters. The clustering first split samples into a group of luminal tumors and a group of TNBCs (with the exception of the ER weakly positive (+25%) luminal tumor PT5_L clustering into the TNBC group). The group of luminal tumors was globally enriched in genes from cluster 1 (early and late response to estrogens); and a subset of luminal tumors were also enriched in genes from the cluster 2 (*TNF* signaling, myogenesis, epithelial mesenchymal transition). The majority of TNBC samples were enriched in genes from cluster 3 (G2M checkpoints, E2F targets, cellular cycle). Genes from cluster 4 showed no clear enrichment in specific pathways and were expressed in complex patterns by both luminal and TNBC tumor subsets. Within each subgroup, the PT samples consistently clustered with their related RD rather than the tumor from the contralateral side. This suggests that beyond the well-known distinct gene expression profiles according to BC subtype, PT and RD are closer from a transcriptomic point of view than are left and right tumors from the same patient.

##### Immune infiltration

We applied the CIBERSORT algorithm to deconvolute RNAseq expression profiles into 22 subsets of immune subpopulations. Out of the 20 samples, the top 3 most abundant immune subpopulations were M2 macrophages (median: 32), CD4 memory resting T cells (median: 26) and M1 macrophages (median: 11), while the majority of immune subpopulations was not identified at significant levels, whether in PT or RD (FigS6A). CD4 memory T cells and M2 macrophages were increased in RD compared with PT (FigS6B); T follicular helper cells and M1 macrophages were significantly higher in TNBC than in luminal BCs, while resting mast cell levels were lower (FigS6C). At the patient level, we did not identify any additional patterns between the left and the paired right tumor (FigS7), nor between the PT and the paired corresponding residual disease (FigS8).

We compared the predicted immune contexture patterns between samples of the cohort using a dissimilarity index (FigS9). Analyzing the entire cohort, the dissimilarity was lower among the PT samples (blue area) than among the RD samples (orange area) (mean: 0.24 *versus* 0.37, *p*=0.009), while the greatest difference was seen between PTs samples compared with RDs samples (yellow area, mean = 0.49)

Analyzing paired data, the mean dissimilarity index was significantly lower between samples of the same side of patient 6 than between the remaining samples (red bordered squares, mean = 0.025), while the dissimilarity indices between the left and the right sides (green bordered squares, mean = 0.22), and between PT and related RDs (yellow bordered squared, mean = 0.29) were not statistically different from the rest of the samples. These results suggest that the composition of the immune microenvironment is strongly driven by the treatment history (PT or RD), while the similarity between a tumor from one side or its contralateral side, or a given tumor and its paired post-chemotherapy sample is lower.

##### TCR sequencing analysis

To further investigate the T cell response to NAC and to compare infiltrating TCR repertoires in clonally distinct tumors within single patients, we extracted TCR beta CDR3 sequences from the RNAseq data, and 6708 clonotypes were identified in the whole cohort. Overall, 89.4% were unique to individual samples (n=5994) (FigS10A), 10.2% were patient specific but shared by multiple samples (n=682), and only 32 clonotypes (0.5%) were identified in different patients (FigS10B).

150 clonotypes (2.2%) were found in VDJdb, a curated database of T-cell receptor sequences of known antigen specificity (FigS10C). The most common antigens for this subset were CMV and Influenza, but the wide majority of clonotypes were unique to a sample. Among 135 clonotypes found both in PT and paired RD, the read count increased significantly after NAC (*p*=0.005) (Fig6D).

**Fig6:**
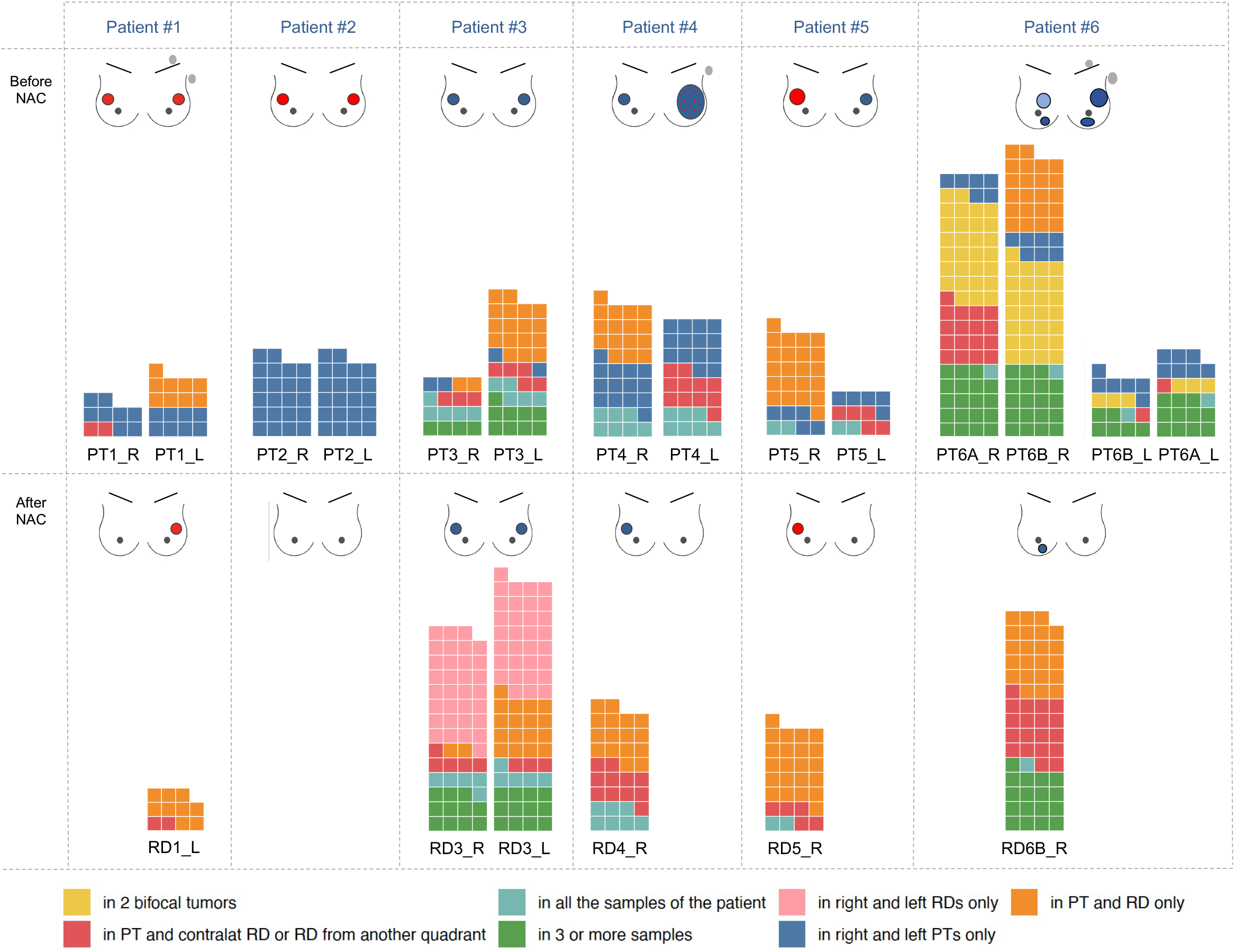
Repartition of the shared clonal TCR repertoire assessed by TCR Beta-chain CDR3 sequences among samples at the individual patient level. The clonotypes are identified with the MixCR algorithm^67^. Only the TCR sequences shared within different samples of a same patient are displayed on the figure. Unique clonotypes (n=5994) and clonotypes shared between patients of the cohort (n=32) are not displayed on the figure. The upper panel represents the repartition of the clonotypes in the 14 PT of the patients #1 to #6, and the lower panel represents the repartition of clonotypes of the 6 samples with RD. Abbreviations: L, left; PT, primary tumor; R, right; RD, residual disease TCR: T cell receptor.

The evolution under NAC of non-unique clones is summarized for each patient in Fig6. While the most common sharing relationships were between PT and matched RD (orange clonotypes) followed by left and right tumor, a subset of clones identified in an index PT were found in the contralateral RD (red clonotypes) or emerged *de novo* in both RD samples of the same patient (pink clonotypes).

Finally, common shared TCRs were extremely rare between individuals, and most of the TCR clonotypes of a tumor were unique. However, a shared TCR repertoire was identified between the left and right primary tumors. An enrichment of TCR repertoire after NAC was identified, including clonotypes deriving a from the contralateral primary tumor.

## Discussion

In the current study, we conducted a large comprehensive overview of sBBCs, integrating clinical, pathological data and data on immune infiltration, together with genomic data generated using modern whole exome and RNA sequencing technologies. It provides important insights to understand the relationships between tumor, host, immunity and response to treatment.

First, we found no indication that sBBCs could be genomically related but rather we conclude that they represent two distinct and independent diseases occurring incidentally at the same time.

In line with previous studies ^28,29,30,31^, we found a high concordance between the clinical and biological patterns within pairs of sBBCs. Notably, the proportion of luminal tumors was very high ^32 33^, while *HER2*-positive BCs were rare as previously reported ^30^. In our study, despite the frequent clinical and pathological concordance between the left and right tumors, we demonstrate clearly that these tumors were genomically independent in terms of mutations, copy number alterations, expression patterns, and clonal composition. This finding is in line with most published studies^8,12,14,19^, despite with older technologies. In line with previous works, we also identified genomically related profiles in multicentric tumors, thus confirming with modern technologies that multi-focal tumors often represent intramammary dissemination of a single breast cancer ^15 14, 17,34^. Overall, these results are consistent with the fact that the occurrence of sBBCs is explained by nongenetic factors ^35^.

Second, we found that the immune infiltration was not purely determined by local tumor microenvironment properties, as TILs levels were different according to the BC subtype of the contralateral tumor. Published studies regarding immune infiltration in sBBCs is scarce, if any. Analyzing samples of sBBCs from 23 patients, one study found that nuclear grade was significantly associated with the number of FoxP3□positive TILs ^36^. In the current work, we analyzed immune infiltration on a large number of sBBCs and confirmed that BC subtype was a master driver of immunosurveillance in BCs. We also found that immune infiltration was modified by the presence of a contralateral tumor: luminal BCs associated with a luminal contralateral BCs had lower str TIL levels than luminal BCs associated with a contralateral BCs of a different subtype. Similarly, a TNBC associated with a contralateral TNBC had higher IT TIL levels than TNBCs associated with a contralateral BC of a different subtype. Several hypotheses can be drawn to explain this observation. First, the immune system might be activated by an index tumor, and immune cells activated by this process might spread to the contralateral tumor; resulting in differences in TIL levels. Second, as luminal BCs associated with a contralateral tumor of another subtype were associated with a lower degree of “luminalness” (as defined by ER and PR staining levels), we cannot exclude that the highest immune infiltration is derived from such patterns rather than from the presence of the contralateral tumor.

Third, the response to NAC was also significantly modified in the case of discordant contralateral tumor subtypes, as with TIL levels. Evidence regarding the influence of contralateral tumor on the response to treatment has been rarely described. In a cohort of 119 patients enrolled in four German neoadjuvant trials, Reinisch and colleagues ^37^ reported that patients with BBCs had lower pCR rates than patients with unilateral BC (12.6% versus 20.9%). These lower rates could partially be explained by the tumor characteristics, as the percentage of hormone receptor positive tumors was significantly higher in the indicator lesions of the sBBCs when compared with the unilateral cancer cohort. However, the response to NAC in sBBCs was lower after statistical adjustment for baseline parameters (*p=* 0.077 with a small number of pCR (13/119)). In our study, it remains unknown whether the difference of the rates of pCR according to the contralateral tumor are the consequence of different baseline immune infiltration levels, or if the contralateral tumor modifies *per se* the chemosensitivity of an index tumor.

Fourth, in tumors resistant to treatment and for which we could perform NGS analysis, the primary tumor and the residual specimen shared a large majority of SNVs and copy number aberrations, but the genomic profile did reveal important changes after NAC. Notably, resistant clones emerged and new potential driver mutations appeared in an independent manner on both sides. Hence, beyond a weak influence of the contralateral tumor on either immune infiltration and/or response to NAC, clonal evolution under NAC seems specific to each tumor.

Finally, we report that the infiltration of T cell clones during NAC leads to an enrichment in the TCR repertoire of post-NAC samples, either derived from the contralateral tumor, either arising in both bilateral tumors with residual disease. At a time where bilateral tumor contexts represent a model of growing interest to understand mechanisms underlying immune response to anticancer treatment in mice ^31, 38^, we provide human data regarding the temporal analysis of the TCR repertoire in sBBCs.

Our study has several strengths. We analyzed genomic and transcriptomic data with modern technologies. Whole exome sequencing is more informative than targeted sequencing for determining clonal relationships, as it enables the comprehensive identification of mutations present in both breast cancers in genomic locations that were not included in the sequencing panel. Whole genome sequencing could further identify mutations in non-coding regions of the genome. Second, we studied a rare and unique cohort of patients, enabling direct comparison of left *versus* right PT together with a temporal analysis comparing paired samples before *versus* after NAC. Hence, beyond all the challenges in analyzing tumor evolution from bulk sequencing data ^39^, we were able to leverage multiple tumor samples to reconstruct a clonal phylogeny from germline data to left and right sBBCs both before and after treatment. Third, to our knowledge, our data on immune infiltration are novel contributions to the literature and provide insights into the immune mechanisms underlying the biology of sBBCs. This study also has limitations. First, we were only able to sequence a limited number of cases. We cannot eliminate the possibility that a subset of clonally related sBBCs could be identified if larger cohorts were sequenced. Second, the cohort was enriched in patients with *BRCA* mutations (although this was not an inclusion criteria), and the latter might represent tumors with particular biological patterns. Recent studies suggest however that the proportion of BBC patients harboring germline pathogenic variants in cancer susceptibility genes may represent up to one third of the patients ^40^. Finally, characterization of the immune microenvironment by bulk sequencing approaches has inherent limitations and new insights could be generated by single cell technologies.

To conclude, our data suggest that the similarity of molecular portraits in sBBCs derives from common environmental factors but not from genetic clonal alterations. Both tumor immune infiltration and response to treatment are influenced by the presence of a contralateral tumor. Pairs of tumors from different BC subtypes should be considered as singular entities before primary systemic treatment is considered as observed responses might deviate from well-known profiles of response to chemotherapy.

## Material and methods

### Patients and treatments

We identified a cohort of 17575 patients with non-metastatic BC treated at the Institut Curie (Paris and Saint Cloud, France) between 2005 and 2015 in the institutional database (CNIL number 1766392 - v1). Patients were treated according to local guidelines. When indicated, chemotherapy was administered in a neoadjuvant or adjuvant setting, endocrine therapy was indicated in the case of positivity for hormone receptor and according to prognostic factors, and patients with *HER2*-positive tumors received neoadjuvant and/or adjuvant trastuzumab from 2007 onwards. This study was approved by the Breast Cancer Study Group and by the Institutional Review Board of Institut Curie and was conducted in accordance with institutional and ethical rules regarding research on tissue specimens and patients. Synchronous bilateral breast cancers (sBBCs) were defined as the occurrence of primary tumors occurring in both breasts with a time interval no greater than 6 months. Metachronous BCs defined as a time interval greater than 6 months between the diagnoses of the first and second tumours were not included in the current study. Patients with exclusive in situ carcinoma (DCIS) in one of the two sides were excluded from the analyses.

### Tumor samples and BC subtype

In accordance with guidelines used in France ^41^, cases were considered estrogen receptor (ER) or progesterone receptor (PR) positive if at least 10% of the tumor cells expressed estrogen and/or progesterone receptors (ER/PR). *HER2* expression was determined by immunohistochemistry with scoring in accordance with American Society of Clinical Oncology (ASCO)/College of American Pathologists (CAP) guidelines^42^. Scores 3+ were reported as positive, score 1+/0 as negative (-). Tumors with scores 2+ were further tested by FISH. *HER2* gene amplification was defined in accordance with ASCO/CAP guidelines. BC subtypes were defined as follows: tumors positive for either ER or PR, and negative for *HER2* were classified as luminal; tumors positive for *HER2* were considered to be HER2-positive BC; tumors negative for ER, PR, and *HER2* were considered to be triple-negative breast cancers (TNBC).

### Pathological review

Bulk tumor specimens - and the corresponding pretreatment core needle biopsy specimens in case of neoadjuvant treatment - were reviewed by an expert in breast pathology (ML). All tumoral tissues studied were Formalin-Fixed Paraffin-Embedded (FFPE) tissue samples. In the cases with bilateral invasive carcinoma, tumor infiltrating lymphocytes (TILs) were reviewed specifically for the purposes of this study, between September 2016 to March 2017. In accordance with the recommendations of the international TILs Working Group^43^, we checked for presence of a mononuclear cell infiltrate in the stroma on hematoxylin and eosin-stained sections without additional staining, after excluding areas around ductal carcinomas *in situ* (DCIS), and tumor zones with necrosis and artifacts. Infiltrates were scored on a continuous scale, as the mean percentage of the stromal area occupied by mononuclear cells. Intratumoral TILs (IT TILs) were defined as intraepithelial mononuclear cells within tumor nests or in direct contact with tumor cells and stromal TILs (Str TILs) were defined as mononuclear inflammatory cells within intratumoral stromal area and were reported as percentage of stromal area. After NAC, we assessed TIL levels within the borders of the residual tumor bed, as defined by the RCB and recommended by the TILs working group ^44^. We simultaneously determined the RCB index, as described by Symmans index ^45^, with the web-based calculator freely available via the Internet (www.mdanderson.org/breastcancer_RCB). We defined pathological complete response (pCR) as the absence of invasive residual tumor from both the breast and axillary nodes (ypT0/is N0).

### Concordance between the tumors of sBBCs pairs

We evaluated the concordance of the clinical and pathological characteristics between the two tumors within the same patient. Pairs of sBBCs composed of tumors of the same BC subtypes were classified as concordant and where otherwise classified as discordant.

### Sample’s preparation and next generation sequencing (NGS) analyses

DNA and RNA were obtained from the Biological Resource Center (BRC of Institut Curie). After selecting patients treated with NAC, tumors from whom sufficient frozen material from both left and right tumors was available in the institutional tissue bank both before (defined as primary tumor (PT)) and after treatment in case of residual disease (RD) were included. Fresh-frozen samples were subjected to genomic DNA extraction and DNA qualification using QuBit system.

#### Whole exome sequencing

1µg of genomic DNA from each sample were subjected to shearing using Covaris system and Illumina compatible libraries were performed according to Agilent SureSelect XT2 library protocol consisting in repairing DNA ends and ligating Illumina barcoded adapters followed by a PCR amplification. Libraries were pooled in equimolar condition before being hybridized on dedicated biotinylated RNA probes targeting whole exome sequences (Agilent Human all Exon V5 capture probes). After selection using streptavidin beads and PCR amplification, enriched library pools were subjected to qPCR quantification using the KAPA library quantification kit (Roche). Sequencing was carried out on the HiSeq2000 instrument from Illumina based on a 2 × 100 cycles mode (paired-end reads, 100 bases) using high output flow cells to produce over 50 and 170 million paired-end reads for 30X (germline) and 100X (tumors) respectively. Samples were sequenced to a median depth of coverage of 153 reads, with 95 % of exonic bases passing 50×coverage. 2 samples were discarded from the analysis due to low purity (RD4_R and RD6B_R). Reads were aligned on the human genome reference hg19/GRCh37 by Burrows-Wheeler Aligner ^46^v0.7.5a; filtering of reads based on target intersection, mapping quality and PCR duplicates removal, using Picard^47^, Bedtools^48^ and Samtools^49^, and preprocess using GATK^50^ for local realignment around indels, and base score recalibration. Preliminary variant calling was performed using Mutect2^51^ for tumor samples, and haplotype caller^52^ for normal samples. SuperFreq v. 1.3.2^53^ performed annotation and filtering of somatic indels and SNVs, copy number and purity estimation, and subclonal reconstruction. SuperFreq was run to analyze together the samples of the same side for each patient. We performed an additional filtering of alterations present in either dbSNP^54^ or ExAC^55^ at a frequency greater than 0.2 or after manual review of the alignment on the Integrative Genomics Viewer (IGV)^56^ or a Somatic Score computed by SuperFreq greater than 0.5. Due to insufficient purity, 2 samples were discarded from the analyses (RD4_R and RD6B_R).

#### Somatic mutations interpretation

Somatic variants were annotated using VEP (version 104) ^57^. A variant was denoted as driver if the mutation was present as either as a splice site, a nonsense, a frameshift or a non-synonymous SNV or indel in COSMIC census gene^58^. The percentage of shared mutations in pairs (pair left right; pair PT-RD; pair of two multicentric tumors of the same side) was defined as the intersect between the mutations present in both tumors *2 divided by the sum of the mutation in the two tumors of the pair *100.

#### Mutational signature deconvolution

The contribution of mutational signatures to individual tumor samples were explored using the signatures deconvoluted by Alexandrov *et al*. ^27^ and referenced in the Cosmic database ^59^. We restricted the analyses to the 13 signatures previously evidenced in BC. Signature activities were estimated using the decompTumor2Sig algorithm ^60^ in the musicatk (version 1.0.0) R package^61^. The percentage of mutational signatures was calculated by summing the relative contribution of each signature in PT samples to the whole tumor spectrum, divided by the number of sampled and the result was multiplied by 100. To avoid overrepresentation of the patient #6 to the cohort and because signatures profiles were similar on each side, we averaged the values of the left side on the one hand, and the values of the right side on the other hand.

#### RNA sequencing

Total RNA extracts from tumor samples were subjected to quality control on a Bioanalyzer instrument and only RNA with RIN (RNA Integrity Number) > 7 were used for sequencing. RNA quantification was achieved using absorbance at 260nm with a Nanodrop spectrophotometer. RNA sequencing libraries were prepared from 1 µg of total RNA using the Illumina TruSeq Stranded mRNA Library preparation kit (following provider’s instructions) which allows performing a strand-specific sequencing. Briefly, a first step of polyA+ RNA selection using oligodT magnetic beads is done to focus sequencing on polyadenylated transcripts. After fragmentation, cDNA synthesis was performed and resulting fragments were used for dA-tailing and then ligated to the TruSeq indexed adapters. PCR amplification is finally achieved to create the final cDNA library. After qPCR quantification using the KAPA library quantification kit (Roche), sequencing was carried out using 2 × 100 cycles (paired-end reads 100 bases). Sequencing was performed by multiplexing barcoded libraries on with the Illumina HiSeq2000 instrument using high output flow cells to obtain 100 million paired-end reads per sample. Alignments were performed on human reference sequences using TopHat^62^ v.2.0.621. Reads with mapping quality <20 and reads marked as duplicates by Picard v.1.97 were excluded from further analysis. Gene-level read counts were obtained using HTSeq-count^63^ and RefSeq hg19/GRCh37.

#### Selection of the genes with the most variant expression and clustering

We selected the most variant genes, based on the inflection point of the interquartile range (IQR) distribution for gene expression. The gene expression was previously rlog transformed with DESeq2^64^. This method is more data-driven than a fixed threshold to define the proportion of genes with the highest level of variation. For each gene, we applied the following procedure: (1) we calculated the IQR for all samples; (2) we sorted the IQR values of the genes in ascending order, to generate an ordered distribution; (3) we estimated the major inflection point of the IQR curve as the point on the curve furthest away from a line drawn between the start and end points of the distribution; (4) we retained genes with an IQR higher than the inflection point. Hierarchical clustering was performed with Pearson distance and Ward linkage.

#### Statistical analysis

The study population was described in terms of frequencies for qualitative variables, or medians and associated ranges for quantitative variables. Chi-squared tests were performed to search for differences between subgroups for each variable (considered significant for *p*-values ≤ 0.05). Continuous variables were compared between groups in Wilcoxon–Mann– Whitney tests for groups of fewer than 30 patients and for variables following multimodal distributions. Student’s t-tests were used in all other cases. Pre and post-NAC TIL levels were analyzed as continuous variables. In case of categorical variables, the kappa coefficient ^65^ was computed as a measure of concordance between the left and the right side of a same patient (varying from - 1 as absolute discordance, to + 1 as absolute concordance, 0 as absence of concordance); otherwise in case of numeric or integer variables, the kendall test was used.

All *P*-values no greater than 0.05 were considered significant. In the case where we tested the hypothesis of potentially different effects of the concordant or discordant status of the tumor pair regarding BC subtype on immune infiltration or response to treatment, we included an interaction term in a linear regression model or logistic regression model respectively. A *p*-value of 0.10 was selected to determine the statistical significance of the interaction term, as it has been suggested due to a low power of the test in the interaction setting ^66^. When appliable, all statistical tests were two-sided. In boxplots, lower and upper bars represent the first and third quartile, respectively, the medium bar is the median, and whiskers extend to 1.5 times the inter-quartile range. Data were processed and statistical analyses were carried out with R software version 3.1.2 (www.cran.r-project.org).

#### Data availability statement

The genomic and transcriptomic data generated during the current study are available in the European genome phenome archive repository, [PERSISTENT WEB LINK TO DATASETS].

#### CIBERSORT

CIBERSORT is an analytical tool quantifying the levels of distinct immune cell types within a complex gene expression mixture (https://cibersort-stanford-edu.proxy.insermbiblio.inist.fr). We applied the original CIBERSORT gene signature LM22 defining 22 immune cell subtypes to all the samples of the cohort, the number of permutations being set to 100, and the mode being set to “absolute”. For each immune subpopulation, (i) we calculated the difference between a given sample and the rest of the cohort; (ii) we squared the result; (iii) and we summed the difference between this patient and each other sample of the cohort, resulting in a dissimilarity index. We displayed the overall results on a correlogram.

#### TCR-sequencing analysis

We applied the MixCR algorithm on RNASeq data of 20 samples to identify and quantify TCR Beta-chain CDR3 sequences. MiXCR (version 2.1.5) ^67^was used with its default parameters to extract and quantify TCR b-chain CDR3 sequences from RNA-seq fastq files. From the MiXCR output, we obtained for each sample the total number of distinct TCRb clones and the number of reads supporting each clone, and we normalized the result by the total number of reads. We investigated if TCR sequences identified by MixCR were identified in the VDJDB database, a curated database of T-cell receptor sequences of known antigen specificity^68^ (Details available at the following URL : https://github.com/antigenomics/vdjdb-db)

## Data Availability

All data produced in the present study are available upon reasonable request to the authors

## Acknowledgments

We thank Corinne Azrijov for collecting patient’s consents, Celine Meaudre for the management of the samples, Audrey Rapinat for DNA and RNA extraction, Antoine Mader for help in designing the figures, Lounes Djerroudi for his advice.

## Author’s contributions

FR and ASHP designed and conceptualized the study.

LD, MD, EL and ASHP identified the Curie cohort and the patient tumor samples.

ASHP and LD managed the clinical database, including performing retrospective chart review.

OM, SV, and MA provided patient tumor samples.

ML provided pathology support and performed pathological review and TILs scoring with TL and BG.

FZ, JA and JH performed literature review.

IB, JYP provided scientific advice and expert guidance on tumor biology.

JA, CL, CV, and ASHP performed the whole exome sequencing analyses.

BS and ASHP performed the RNASeq analyses.

WR provided guidance for MixCR analyses.

JW provided scientific direction.

FR and ASHP allocated funding.

All the authors participated in the preparation of the manuscript.

