## Supplemental Material for "Evolution of synchronous bilateral breast cancers provide insights into interactions between host, tumor and immunity"

***Patient #1*** (Fig4A*)* was diagnosed with a bifocal highly proliferative triple negative sBBC with a low bilateral immune infiltration in both sides, in the context of a *BRCA1* germline mutation. After NAC, the right tumor reached pCR, while tumor cells were not modified following NAC in the left side (RD 37 mm, 1 node involved, high post-NAC mitotic index) as determined by histology. Genomic profiling of the 2 PTs identified no shared somatic mutations between the left and the right side. Both PTs were mutated for *P53*, but the genomic alteration was different on the left and the right side (left side: non-synonymous substitution c.524G>A, p.R175H, right side: frameshift deletion c.636delT, p.F212del). Other potential driver alterations included (i) right side: *PML, MLH1, NOTCH2, SETBP1, GNA11*; (ii) left side: 3 mutations shared between PT and RD (*TP53* (clinvar), *CIC*, *ATP23B*); and 3 mutations specific to the RD (*CD40LG* (clinvar); *RB1*; *BCL11B*). Clonality analyses of the right PT-RD pair suggested clonal extinction of one subclone, while 2 subclones emerged and represented the majority of the RD, consistent with the potential driver function of the three RD specific mutations. After bilateral mastectomy, the patient was treated with 5FU based adjuvant chemotherapy but was diagnosed with an early clinical local relapse on the left side nine months later and died 12 months after relapse.

***Patient #2*** (Fig4B) was diagnosed with bilateral highly proliferating TNBC (right: bifocal; left: unifocal) in the context of *BRCA1* germline mutation. At the genomic level, no mutation was shared between both sides. Both tumors were mutated for *TP53* but with different mutations (left side: frameshift deletion c.213delT, p.S241del, right side: frameshift insertion c.498-499insAC, p.S166-Q167X). Four additional potential driver mutations were found in the left side (*KIT*, *PTEN, WHSC1L1, ZMYM2*). Both tumors reached pCR after NAC.

***Patient #3*** (Fig4C) was diagnosed with synchronous bilateral luminal BC (right: luminal B (grade III, KI40%); left luminal A (lobular grade I, ER+90% PR+90%)). Neither of the tumors reached pCR after NAC. At the genomic level, no mutations were shared between both sides. On the right side, a frameshift mutation on *JUN* occurred early and remained in the RD. On the left side, the number of mutations was far higher than in the right tumor (103 and 24 respectively), consistent with the observation that the left side was dominated by a high amount (Ciriello et al., 2015)of signature 3 despite no evidence for mutation or methylation of *BRCA1* nor *RAD51C*. Only one driver gene (*CASC5*) was identified in the PT, and was not observed in the RD. The lobular tumor was mutated for *CDH1* as previously described (substitution T>A p.S18T nonsynonymous substitution).

***Patient #4*** (Fig4D) was diagnosed with sBBC, in the context of *BRCA2* germline mutation (left: locally advanced inflammatory luminal B BC; right: luminal A BC). While the left tumor reached pCR, the right tumor had a high residual burden, with notably extensive lymphovascular invasion and 2 nodes with capsular rupture. At the genomic level, no mutation was shared between left and right tumor. On the right side, five potential driver mutations were found in the PT (*PTEN* (clinvar), *ARID1A*, *STAT5B*, *TRRAP*, *PTPRC*). The low tumor purity of the RD sample precluded WES analysis. On the left side, 4 candidate driver mutations were found (*PIK3CA*, *STAG2*, *ROS1,* *TBX3* and *COL6A3*).

***Patient #5*** (Fig4E) was diagnosed with bilateral discordant BCs (right: TNBC; left: luminal B (grade III, ER+25% PR-, Ki67: 80%)). No mutation was shared between left and right tumor. TP53 was mutated on both sides (left side: frameshift deletion c.636delT, p.F212del, right side: frameshift deletion c.165delT, p.T55del).

On the left side, three potential driver mutations were found (*BRCA1*, *TP53*, *PTEN*). The presence of a *BRCA1* mutation was consistent with (i) a high proportion of signature 3 and (ii) a higher global number of mutations on the left side when compared with the right side (226 and 65 respectively). The tumor reached pCR after NAC. On the right side, three potential driver mutations were identified (*TP53*, *MECOM*, and *RNF43)* and were present in the residual disease, while a mutation in *ARID1A* was specific to PT. The clonality analysis revealed that most of the clones present in RD were already present in the PT, and that the chemotherapy had little impact on clonal evolution.

***Patient #6*** (Fig4F) was diagnosed with bilateral multicentric BBC, in the context of *BRCA1* germline mutation (three tumors luminal B, one tumor luminal A). No mutations were shared between sides but most mutations were shared between the 2 tumor foci of each side. Clonality analyses showed that tumors were clonally related on both sides, though it was not possible to assess which tumor was the ancestor of the other one.

On the left side, common mutations between the two tumors were identified on *CFH*, *PMS1*, *EGFR*, *NONO*, *TRAF7*, *SMARCA4*, while *RAP1GDS1* was specific of tumor B. On the right side, two potential drivers were identified in both tumors (*HOXC11*, *CRTC3*), while *CDC42EP1* was specific of tumor A.
