## Supplemental figures and tables' legends for "Evolution of synchronous bilateral breast cancers provide insights into interactions between host, tumor and immunity"

**FigS1: Flow chart of the study cohort.**

Abbreviations: BC, breast cancer; RNAseq, RNA sequencing; sBBC, synchronous bilateral breast cancers; TIL, tumor infiltrating lymphocytes; WES, whole exome sequencing.

**FigS2: Comparison of the patient's and tumor's characteristics according to the unilateral or bilateral character of the tumor.**

The sBBCs cohort comprises 404 patients carrying 804 sBBCs that are compared with 17171 patients with unilateral BCs. Comparison of the bilaterality regarding (i) patients characteristics: A. Age at BC diagnosis; B. BMI; C. Presence of a germline mutation of *BRCA1* or *BRCA2*; D. First treatment; E. Breast surgery; F. Chemotherapy; G. Endocrine therapy; (ii) tumor’s characteristics: H. ER staining (as percentage); I. PR staining (as percentage); J. BC subtype; K. Histological type; L. Tumor grade; M. Lymphovascular invasion; N. Multifocality.

Abbreviations: BC, breast cancer; BMI, body mass index; ER, estro gen receptor; LVI, lymphovascular invasion; NET, neoadjuvant endocrine therapy; NRT, neoadjuvant radiotherapy; PR, progesterone receptor.

**FigS3: Patient's and tumor's characteristics and response to neoadjuvant chemotherapy in the 6 patients (20 samples) with NGS data.**

Primary tumors are labelled PT, specimens with residual disease are labelled RD.

Abbreviations: ER, estrogen receptor; IT TIL, intra tumoral tumor infiltrating lymphocytes; L, left; neg, negative; pCR, pathologic complete response; PR, progesterone receptor; PT, patient; R, right, RD, residual disease; str TIL, stromal tumor infiltrating lymphocytes; TNBC: triple negative breast cancer.

**FigS4: Copy number patterns of the genomes of the 14 primary tumors.**

The total percentage of gain/loss is listed by the cumulative change per sample.

Chr, chromosome

**FigS5: Transcriptomic analyses of the sBBCs cohort.**

Gene expression clustering with RNA-sequencing data based on the 2846 most variant genes on the in left, right, pre and post-NAC samples of a cohort of 6 patients (20 samples) with RNASeq data available. Cluster 1 is enriched in genes coding for early and late response to estrogens; cluster 2 is enriched in genes coding for TNF signaling, myogenesis, epithelial mesenchymal transition; cluster 3 is enriched in genes coding for G2M checkpoints, E2F targets, cellular cycle; cluster 4 shows no clear enrichment in specific pathways.

Abbreviations: L, left; PT, patient; R, right, RD, residual disease; TNBC: triple negative breast cancer; wt, wild type.

**FigS6: Composition of the immune microenvironment (22 immune subpopulations) of the sBBCs cohort (from the CIBERSORT algorithm).**

The analyses are performed in the in left, right, pre and post-NAC samples of a cohort of 6 patients (20 samples). The composition of the immune microenvironment (22 immune subpopulations) was deconvoluted by the CIBERSORT algorithm (absolute mode) as described by Newman (Newman et al., 2015);

A. Composition of the immune microenvironment in the whole population, in PT, and in RD samples; B Comparison of the levels of the top 12 immune subpopulations gene expression according to the type of sample (PT *versus* RD); C Comparison of the levels of the top 12 immune subpopulations gene expression according to the BC subtype (luminal *versus* TNBC);

Abbreviations: L, left; NAC, neoadjuvant chemotherapy; PT, primary tumor; R, right; RD, residual disease; sBBC, synchronous bilateral breast cancer; TNBC, triple negative breast cancer.

**FigS7: Comparison of the different subsets of immune subpopulation according to specific conditions.**

A. Comparison according to the PT or the RD character of the sample; B. Comparison according to the PT or the RD character of the sample and the achievement of a pCR; C. Comparison according to the BC subtype of the sample. The absolute amount of immune population is calculated using the CIBERSORT algorithm; Immune subpopulations with levels below 0.1 arbitrary unit are discarded from the analyses.

L, left; pCR, pathologic complete response; NAC, neoadjuvant chemotherapy; PT, primary tumor; right, right; RD, residual disease; TNBC, triple negative breast cancer

**FigS8: Comparison of the repartition of the subsets of immune subpopulation in the left and the right tumors of pairs of sBBCs.**

Each subfigure represents the amount of immune subpopulation evaluated with the CIBERSORT algorithm; in the right (R) tumor (left panel), the left (L) tumor (middle panel), and the absolute difference between the left and the right tumor (right panel). Patient 1 to 6 are represented on subfigures A to F. Both tumors of each side of patient 6 are averaged, as they show very minimal differences in the composition of the immune microenvironment.

L, left; R, right.

**FigS9: Comparison of the similarity of the immune microenvironnement across samples.**

The analyses are performed in the in left, right, pre and post-NAC samples of the cohort of 6 patients (20 samples). The composition of the immune microenvironment (22 immune subpopulations) was deconvoluted by the CIBERSORT algorithm (absolute mode) as described by Newman (Newman et al., 2015). A dissimilarity index of the immune microenvironnement was displayed sample *versus* sample. The colored area represents the samples compared at the cohort levels (dissimilarity among the PT samples (blue area), among the RD samples (orange area), and between PTs samples compared with RDs samples (yellow area). The colored bordered squares represent the comparison of paired data (samples of the same side of a patient (red bordered squares), the left and the right sides (green bordered squares), and between PT and related RDs (yellow bordered squared).

L, left; PT, primary tumor; R, right; RD, residual disease;

**FigS10: Repartition of the clonal TCR repertoire assessed by TCR Beta-chain CDR3 sequences at the cohort level.**

The clonotypes are identified with the MixCR algorithm(Bolotin et al., 2015); A. Repartition of the clonotypes in the cohort. Clonotypes that are shared between patients are represented by a dark blue bar, shared within the different samples of a patient are represented by a light blue bar, and singletons (clonotypes occurring once) are represented in grey. B. Repartition of clonotypes shared between patients; C. Repartition of the 150 TCR sequences identified in VDJDB (curated database of T-cell receptor sequences of known antigen specificity); D. Clone dynamics among clonotypes shared between PTs and RDs.

L, left; NAC, neoadjuvant chemotherapy; PT, primary tumor; R, right; RD, residual disease.

**TableS1: Comparison of the patients and tumor characteristics according to the unilateral or synchronous bilateral character of the tumor.**

Missing data: Age, n=2; Age class, n=2; BMI, n=5943; BMI class, n=5943; BMI class, n=5943; Age at menarche, n=7543; Previous pregnancy, n=6239; Menopausal status, n=2828; Age at menopause, n=9428; Hormone replacement therapy, n=12483; Familial history breast / ov. cancer, n=8875; Research hereditary predisposition, n=13725; BRCA mutation, n=16178; Targeted therapy type, n=31; Diagnostic modality, n=5851; clinical size, n=8481; clinical T stage, n=472; clinical N stage, n=29; pathological T stage, n=2594; BC subtype, n=2797; ER status, n=2588; PR status, n=3349; HER2 status, n=2776; percentage of ER positivity, n=2675; percentage of PR positivity, n=3420; Invasive or DCIS, n=2451; DCIS component, n=7418; histological size, n=5221; grade, n=2846; lymphovascular invasion, n=5388; histological type, n=88; axillar surgery, n=805.

Abbreviations: AND, axillary lymph node dissection; BC, breast cancer; BMI, body mass index; DCIS, ductal carcinoma in situ; ER, estrogen receptor; LVI, lymphovascular invasion; NET, neoadjuvant endocrine therapy; NRT, neoadjuvant radiotherapy; NST, no special type; PR, progesterone receptor; SND, sentinel lymph node dissection; TNBC: triple negative breast cancer.

**TableS2: Patient's and tumor characteristics of the cohort of the invasive sBBCs.**

Missing data: Age, n=1; Age class, n=1; BMI, n=18; BMI class, n=18; Previous pregnancy, n=15; Menopausal status, n=13; Familial history breast / ov. cancer, n=74; Research hereditary predisposition, n=223; BRCA mutation, n=263

Diagnostic modality, n=25; clinical size, n=288; clinical T stage, n=5; clinical N stage, n=2; pathological T stage, n=142; BC subtype, n=12; ER status, n=8; PR status, n=29; HER2 status, n=12; intensity of ER positivity, n=77; percentage of ER positivity, n=16; intensity of PR positivity, n=150; percentage of PR positivity, n=37; DCIS component, n=19; pN status, n=139; grade, n=45; lymphovascular invasion, n=156; histological type, n=5; axillar surgery, n=6.

Abbreviations: BC, breast cancer; DCIS, ductal carcinoma in situ; ER, estrogen receptor; NET, neoadjuvant endocrine therapy; NRT, neoadjuvant radiotherapy; NST, no special type; PR, progesterone receptor; SND, sentinel lymph node dissection; TNBC: triple negative breast cancer.

**TableS3: Concordance between left and right tumor's characteristics.**

Abbreviations: BC, breast cancer; BMI, body mass index; DCIS, ductal carcinoma in situ; ER, estrogen receptor; IT TIL, intra tumoral tumor infiltrating lymphocytes; PR, progesterone receptor; str TIL, stromal tumor infiltrating lymphocytes; TNBC: triple negative breast cancer.

**TableS4: Association between patient's tumor's characteristics and immune infiltration (str and IT TIL levels).**

Abbreviations: BC, breast cancer; BMI, body mass index; DCIS, ductal carcinoma in situ; ER, estrogen receptor; NST, no special type; PR, progesterone receptor; TNBC: triple negative breast cancer.

**TableS5: Patient's and tumor's characteristics of the 6 patients and 20 samples with NGS data.**

Abbreviations: AND, axillary lymph node dissection; BC, breast cancer; ER, estrogen receptor; IT TIL, intra tumoral tumor infiltrating lymphocytes; L, left; NAC, neoadjuvant chemotherapy; PR, progesterone receptor; R, right; RCB, residual cancer burden; SND, sentinel lymph node dissection; str TIL, stromal tumor infiltrating lymphocytes; wt, wild type.

**TableS6: Mutations per sample and per patient in the cohort of the 6 patients and 20 samples with NGS data.**

Variant characteristics are derived from SuperFreq results and VEP annotations. sample_mutation_id refers to the name of the sample, the number of the chromosome, followed by the start position of the mutation; Location refers to the number of the chromosome, followed by the start position and the end position of the mutation; severity is a numeric version of the effect which denotes the mutation type; f is the variant allele frequency (i.e. ratio between the number of variant reads and the total number of reads covering the mutation position); cov, ref and var denote the number of total reads, reads with the reference sequence, and reads with the variant respectively; flag reports potential flags from the original mutation calling; pbq and pmq are quality measures for base and mapping; psr quantifies the strand ratio for the reads; SomaticScore and germlineLike are a global scores computed by SuperFreq to assess whether a mutation is germline or somatic; the subsequent columns refer to known variant databases: dbSNP, ExAC, Cosmic and ClinVar; mutation_type specifies the type of mutation (SNV or indel); clone is the id of the clone in which the mutation appeared, according the the evolutionary reconstruction from SuperFreq; ccf is the estimated cancer cell fraction; Allele the variant sequence; and finally VEP annotations corresponding to the first reported transcript.
