## Supplementary figures and images for "Evolution of synchronous bilateral breast cancers provide insights into interactions between host, tumor and immunity"

### Supplemental Figure 1

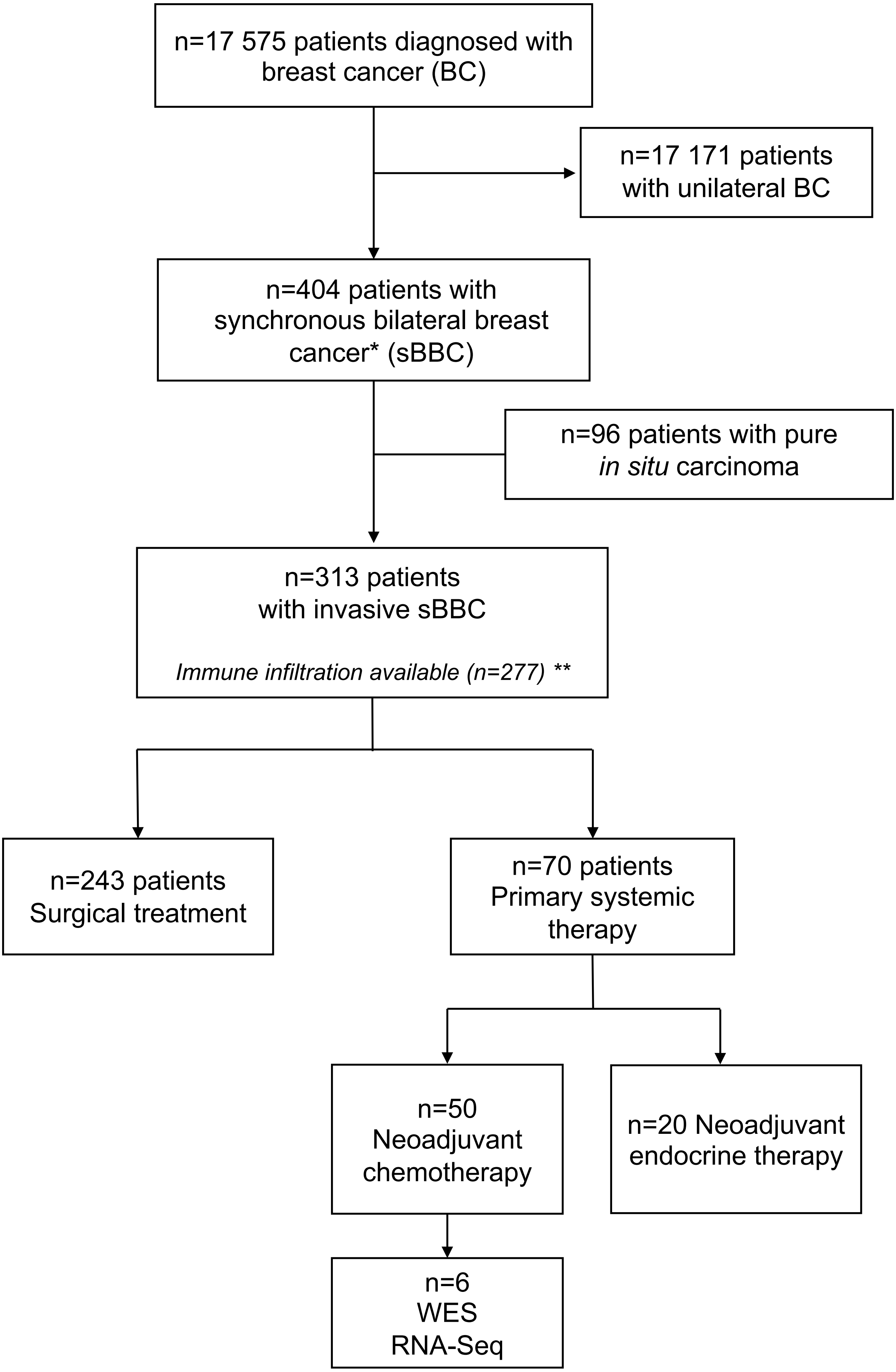


* diagnosed within 6 months after a first breast cancer

* * Stromal TILs (n=277) ; intra tumoral TILs (n=275)

### Supplemental Figure 2

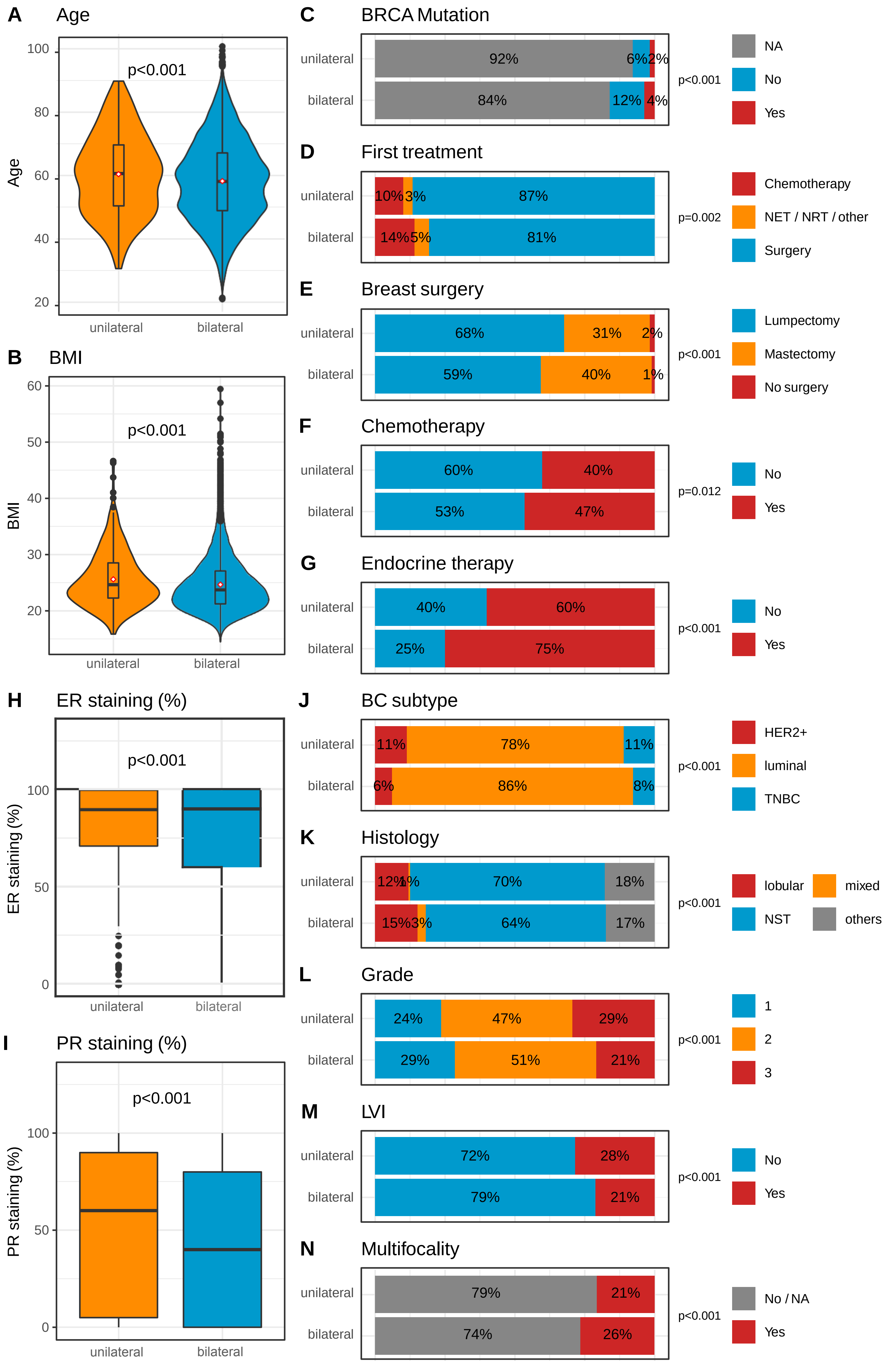

### Supplemental Figure 3

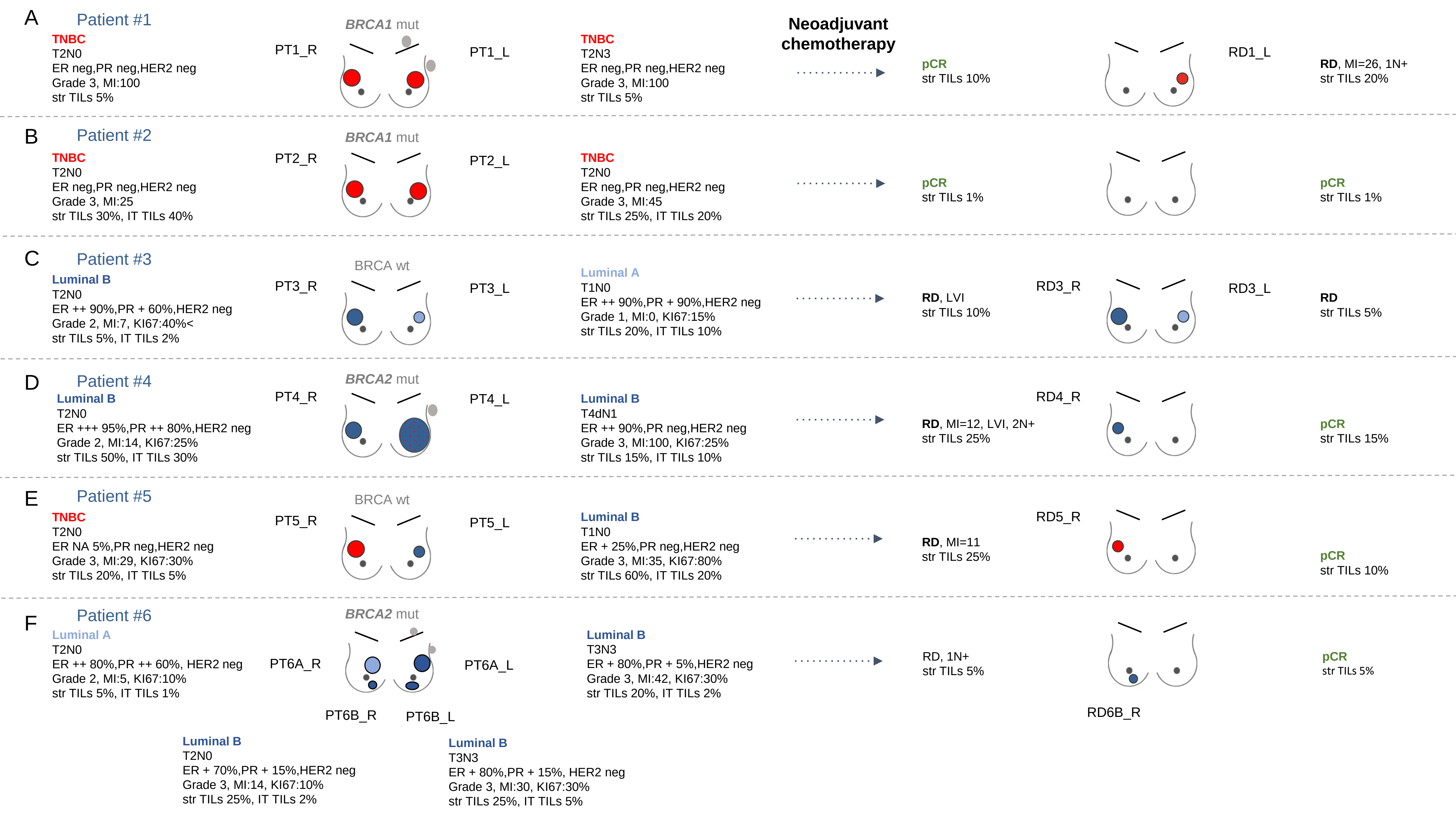

### Supplemental Figure 4

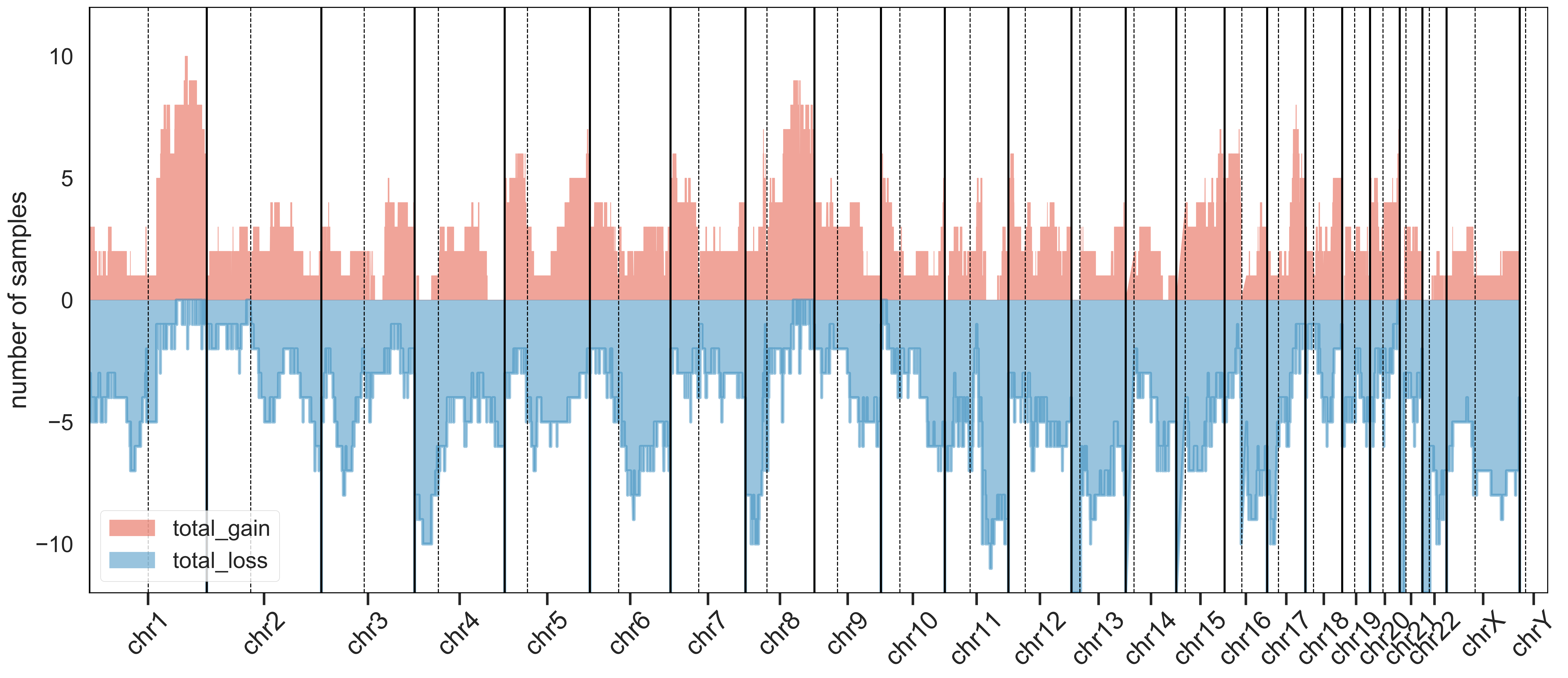

### Supplemental Figure 5

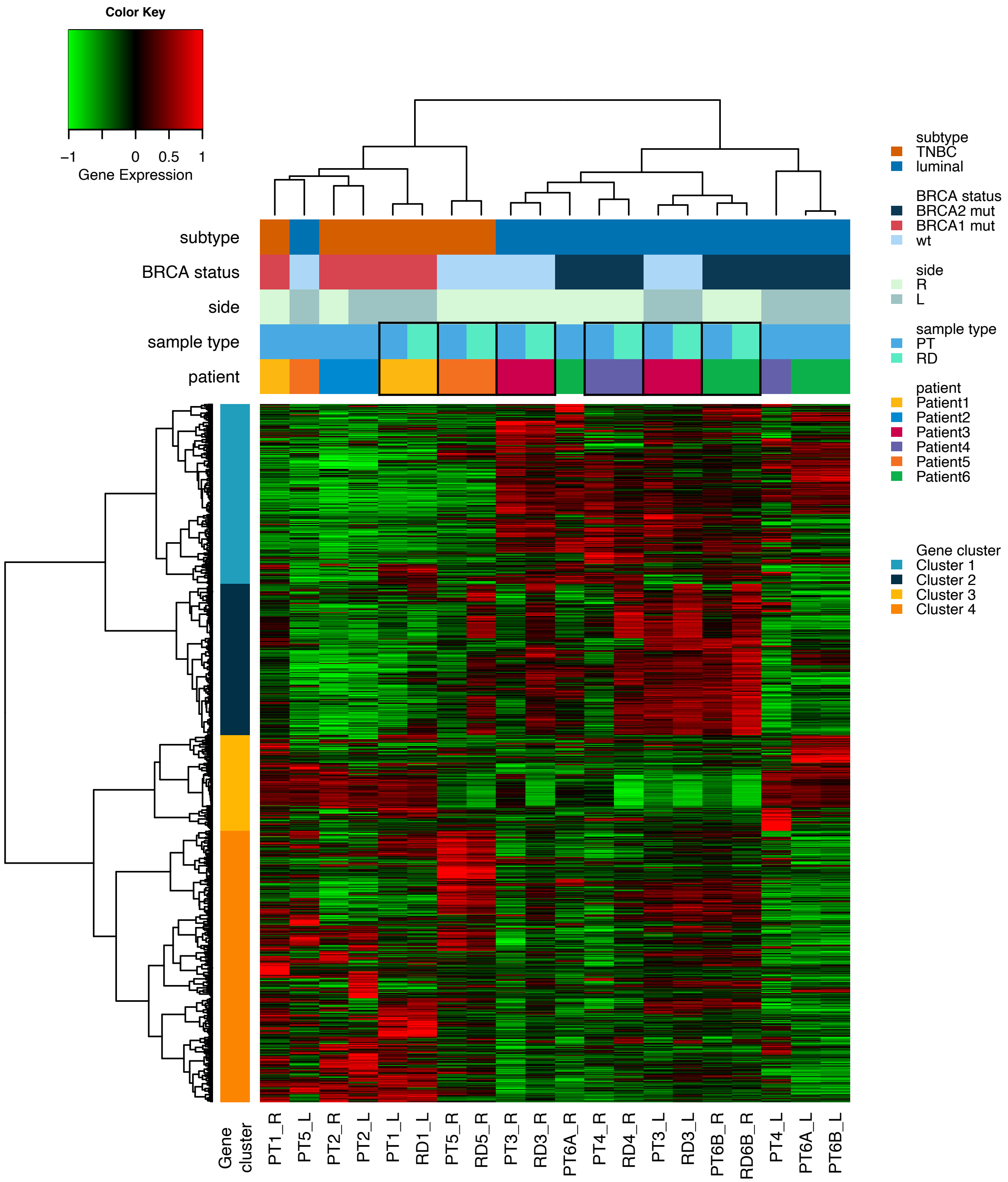

### Supplemental Figure 6

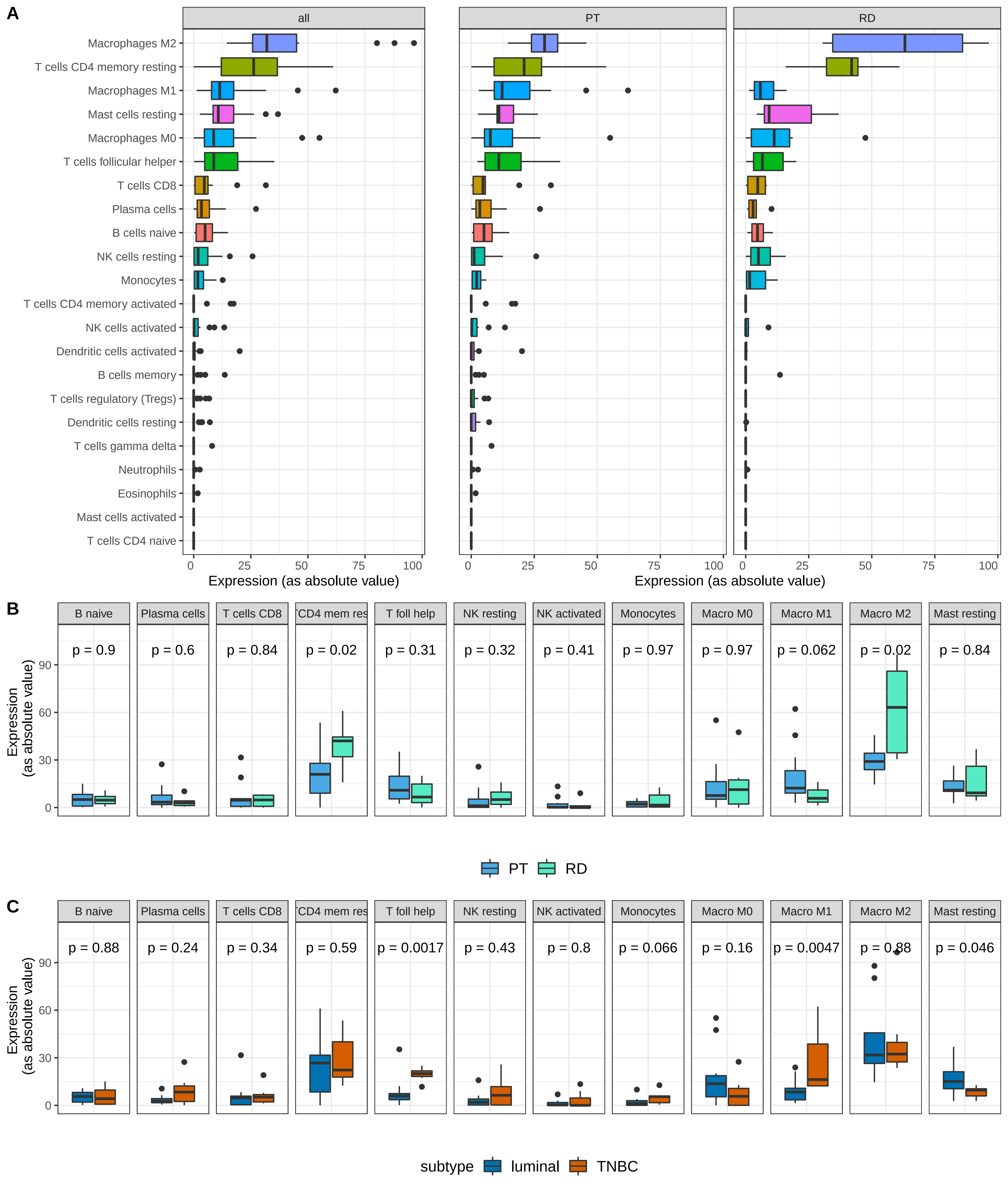

### Supplemental Figure 7

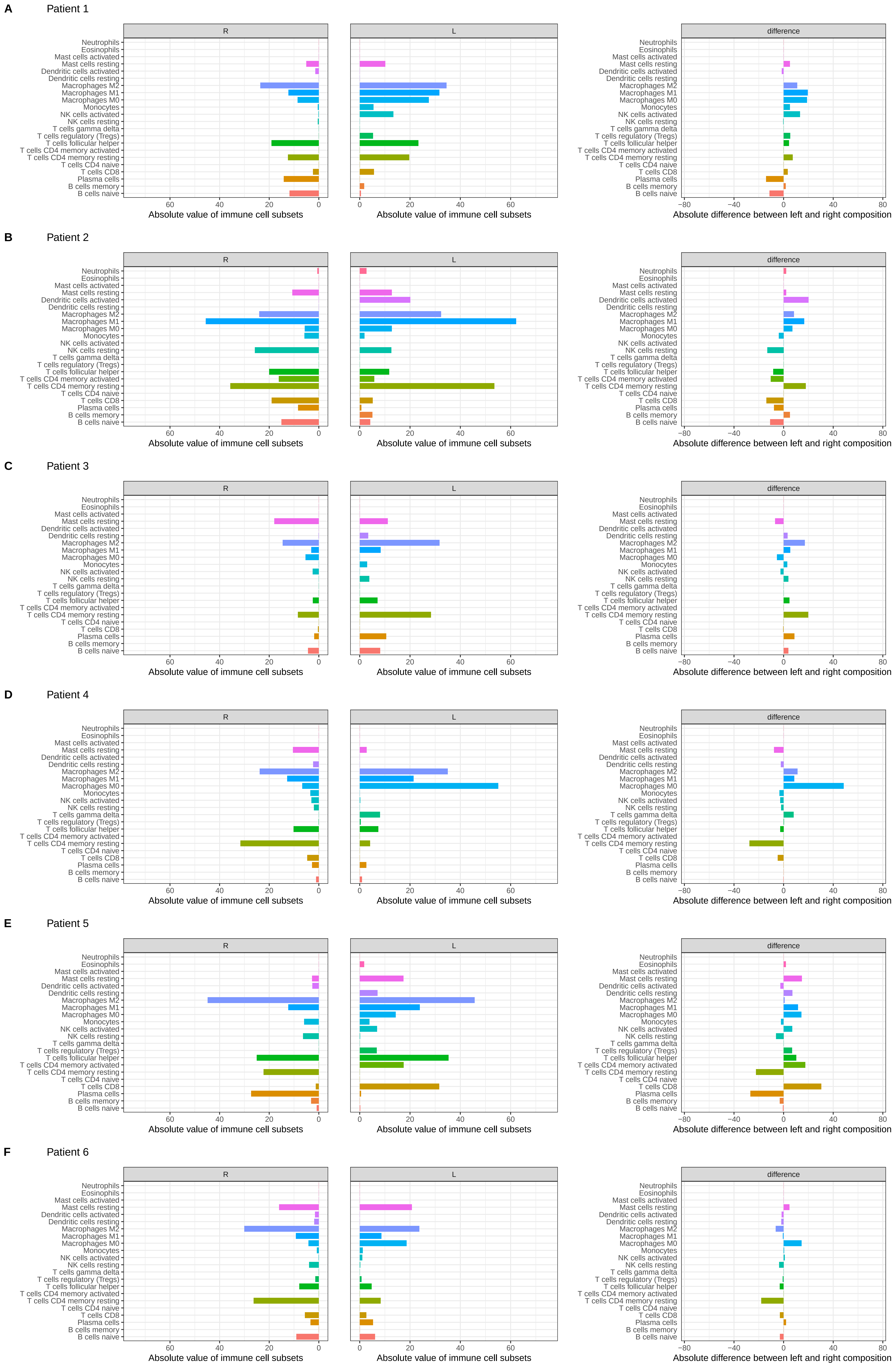

### Supplemental Figure 8

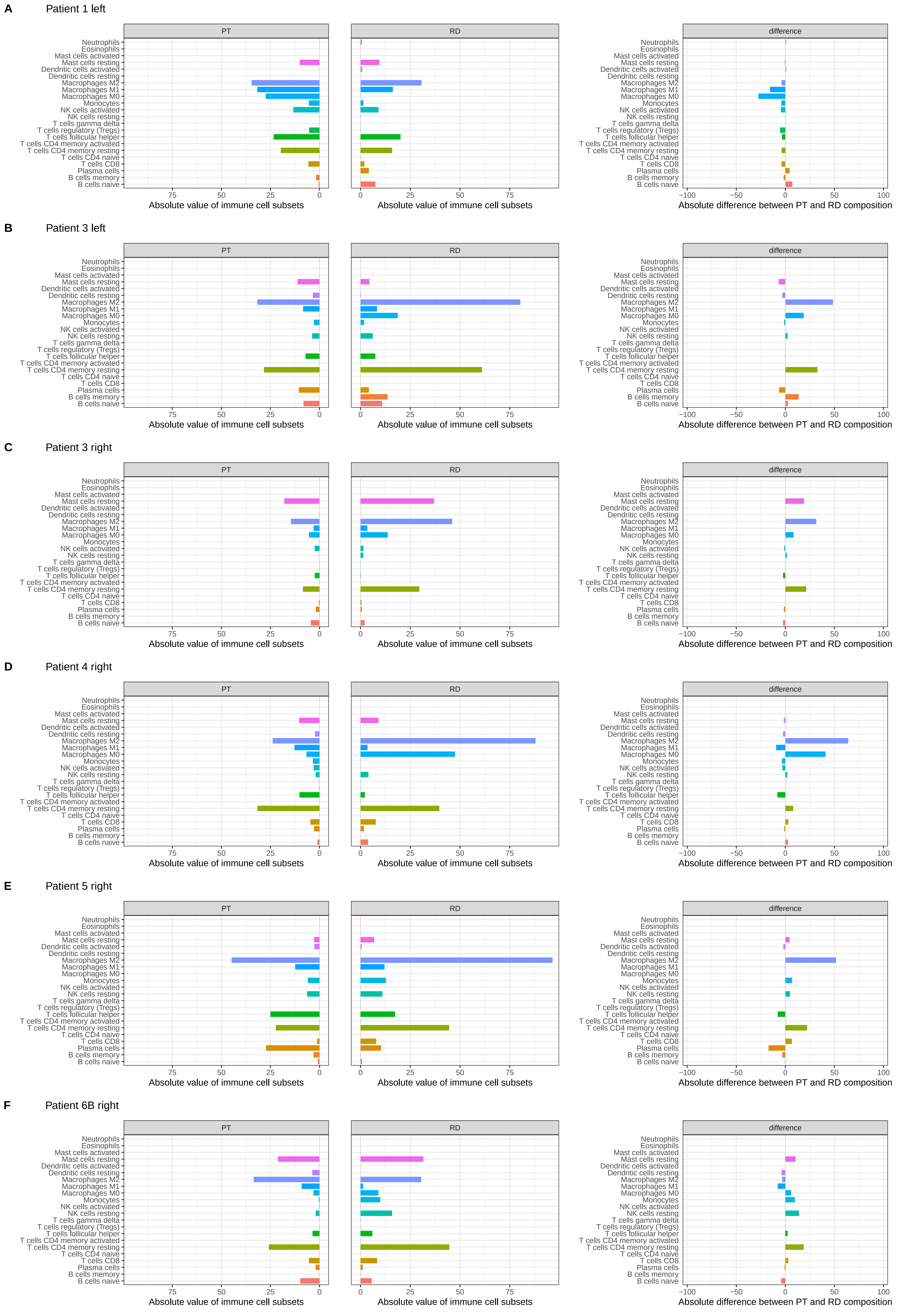

### Supplemental Figure 10

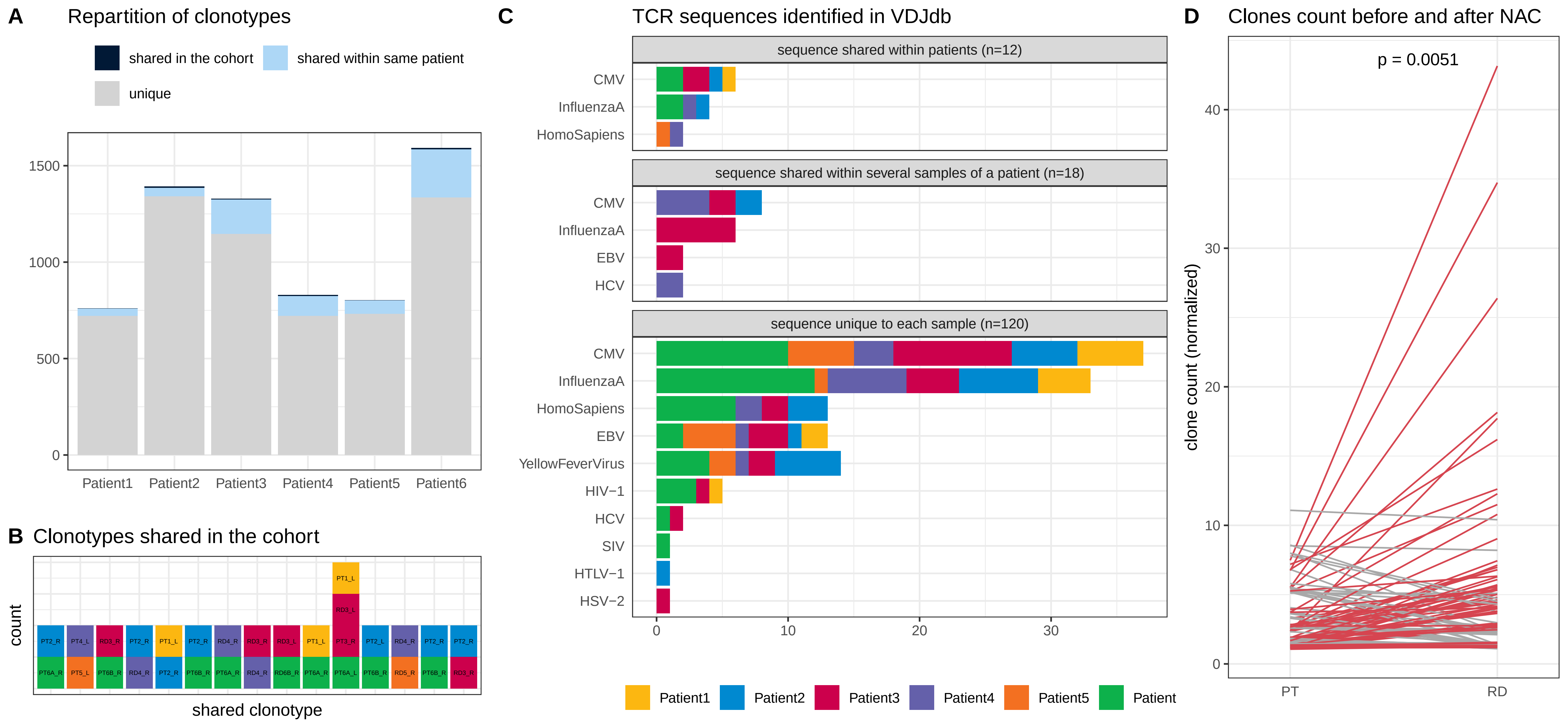
