## Supplemental Figure 9 for "Evolution of synchronous bilateral breast cancers provide insights into interactions between host, tumor and immunity"

| | PT1_L | PT2_R | PT2_L | PT3_R | PT3_L | PT4_R | PT4_L | PT5_R | PT5_L | PT6B_L | PT6A_L | PT6B_R | PT6A_R | RD1_L | RD3_R | RD3_L | RD4_R | RD5_R | RD6B_R |
|---|---|---|---|---|---|---|---|---|---|---|---|---|---|---|---|---|---|---|---|
| PT1_R | 0.15 | 0.3 | 0.52 | 0.09 | 0.06 | 0.08 | 0.29 | 0.1 | 0.29 | 0.06 | 0.1 | 0.1 | 0.05 | 0.04 | 0.26 | 0.61 | 0.7 | 0.68 | 0.26 |
| PT1_L | | 0.26 | 0.33 | 0.26 | 0.21 | 0.14 | 0.18 | 0.23 | 0.2 | 0.18 | 0.15 | 0.2 | 0.16 | 0.12 | 0.27 | 0.53 | 0.52 | 0.61 | 0.32 |
| PT2_R | | | 0.19 | 0.45 | 0.29 | 0.25 | 0.6 | 0.34 | 0.38 | 0.42 | 0.38 | 0.3 | 0.24 | 0.27 | 0.49 | 0.68 | 0.9 | 0.74 | 0.34 |
| PT2_L | | | | 0.67 | 0.44 | 0.37 | 0.68 | 0.52 | 0.67 | 0.63 | 0.54 | 0.45 | 0.41 | 0.45 | 0.56 | 0.6 | 0.86 | 0.76 | 0.47 |
| PT3_R | | | | | 0.09 | 0.09 | 0.36 | 0.27 | 0.41 | 0.02 | 0.05 | 0.08 | 0.08 | 0.1 | 0.19 | 0.78 | 0.83 | 0.89 | 0.22 |
| PT3_L | | | | | | 0.03 | 0.41 | 0.1 | 0.39 | 0.1 | 0.11 | 0.02 | 0.02 | 0.05 | 0.13 | 0.41 | 0.58 | 0.49 | 0.12 |
| PT4_R | | | | | | | 0.35 | 0.15 | 0.35 | 0.1 | 0.09 | 0.04 | 0.02 | 0.06 | 0.15 | 0.46 | 0.6 | 0.58 | 0.11 |
| PT4_L | | | | | | | | 0.46 | 0.43 | 0.21 | 0.21 | 0.39 | 0.35 | 0.36 | 0.41 | 0.72 | 0.46 | 0.91 | 0.55 |
| PT5_R | | | | | | | | | 0.33 | 0.23 | 0.25 | 0.18 | 0.12 | 0.11 | 0.29 | 0.42 | 0.58 | 0.36 | 0.3 |
| PT5_L | | | | | | | | | | 0.34 | 0.29 | 0.34 | 0.32 | 0.24 | 0.43 | 0.79 | 0.71 | 0.66 | 0.51 |
| PT6B_L | | | | | | | | | | | 0.02 | 0.09 | 0.08 | 0.09 | 0.18 | 0.7 | 0.67 | 0.81 | 0.24 |
| PT6A_L | | | | | | | | | | | | 0.06 | 0.08 | 0.11 | 0.1 | 0.62 | 0.57 | 0.73 | 0.17 |
| PT6B_R | | | | | | | | | | | | | 0.03 | 0.07 | 0.07 | 0.42 | 0.54 | 0.51 | 0.09 |
| PT6A_R | | | | | | | | | | | | | | 0.04 | 0.14 | 0.46 | 0.59 | 0.56 | 0.11 |
| RD1_L | | | | | | | | | | | | | | | 0.2 | 0.54 | 0.67 | 0.56 | 0.23 |
| RD3_R | | | | | | | | | | | | | | | | 0.36 | 0.38 | 0.46 | 0.09 |
| RD3_L | | | | | | | | | | | | | | | | | 0.17 | 0.15 | 0.41 |
| RD4_R | | | | | | | | | | | | | | | | | | 0.3 | 0.56 |
| RD5_R | | | | | | | | | | | | | | | | | | | 0.54 |
